# Elevated T-cell exhaustion and urinary tumor DNA levels are associated with BCG failure in patients with non-muscle invasive bladder cancer

**DOI:** 10.1101/2022.03.23.22272806

**Authors:** Trine Strandgaard, Sia Viborg Lindskrog, Iver Nordentoft, Emil Christensen, Karin Birkenkamp-Demtröder, Tine Ginnerup Andreasen, Philippe Lamy, Asbjørn Kjær, Daniel Ranti, Yuan-Sho Wang, Christine Bieber, Frederik Prip, Julie Rasmussen, Torben Steiniche, Nicolai Birkbak, John Sfakianos, Amir Horowitz, Jørgen Bjerggaard Jensen, Lars Dyrskjøt

**Author notes:** **Corresponding author:** Lars Dyrskjøt.

## Abstract

**Background:** The functional status of immune cells within the tumor microenvironment and tumor characteristics may explain Bacillus Calmette-Guérin (BCG)-failure in high-risk non-muscle invasive bladder cancer (NMIBC).

**Objective:** To characterize molecular correlates of BCG-failure using a multiomics approach.

**Design, Setting, and Participants:** BCG-treated NMIBC patients (n=156) were included. Metachronous tumors were analyzed using RNA-sequencing (n=170) and whole exome sequencing (n=198). Urine samples were analyzed for immune-oncology related proteins (n=190), and tumor-derived DNA (tdDNA; n=192).

**Outcome Measurement and Statistical Analysis:** Primary endpoint was BCG-failure. Cox regression, Wilcoxon Rank Sum test, t-test or Fisher’s exact test were used.

**Results and Limitations:** BCG caused activation of the immune system regardless of clinical response; however, immune-inhibitory proteins were observed in the urine of BCG-unresponsive patients post-treatment (CD70, PD1, CD5). BCG-failure was associated with post-BCG T-cell exhaustion (*p*=0.0021). Pre-BCG tumors from patients with post-BCG T-cell exhaustion were characterized by high expression of cell division and immune-related genes. A high post-BCG exhaustion prediction score in pre-BCG tumors was associated with worse post-BCG high-grade recurrence free survival (HGRFS), reflecting BCG-failure (*p*=0.0084). Pre-BCG tumors of class 2a and 2b were likewise associated with worse post-BCG HGRFS(*p*=0.0023). Post-BCG exhaustion was observed in patients with high pre-BCG neoantigen load (*p*=0.023) and mutations in *MUC4* (*p*=0.0007). Finally, absence of post-BCG tdDNA clearance identified patients at high risk of recurrence (*p*=0.028). The retrospective design, lack of maintenance BCG, and partial overlap in analyses are limitations to the study.

**Conclusions:** BCG failure may be caused by T-cell exhaustion. Tumor subtype and Pre-BCG tumor characteristics may identify patients at high risk of BCG-failure prior to treatment. Urinary measurements have the potential to be used as a real-time assessment of treatment response.

**Patient Summary:** A dysfunctional immune response to BCG therapy may explain lack of response to the treatment.

## Introduction

Treatment of patients with high risk non-muscle invasive bladder cancer (NMIBC) includes intravesical instillations with the immunotherapy Bacillus Calmette-Guérin (BCG) to reduce the risk of recurrence and progression^1^. Though BCG has been used for decades, the mechanisms of action and failure are not fully understood. Despite of high initial response rates, a study found that up to 42% of patients recurred after complete BCG-therapy, and 14% progressed^2,3^

BCG is thought to act through activation of both the innate and adaptive immune systems, thereby promoting lasting antitumor effects^4^. Several immune cell types have proven necessary for BCG-response *in vitro* and in murine studies, including natural killer (NK) cells^5^, neutrophils^6^, and T-cells^7^. Human studies demonstrating the important immune cellular mechanisms associated with response and resistance are still scarce. However, the composition of the tumor microenvironment (TME) has been shown to be important for disease outcome as well as response to treatment in both bladder cancer and other cancers^8,9^. Additionally, patients with increased levels of Type 1 T-helper cell (T_H_1) cytokines during and after treatment^10–12^ and increased pre-treatment numbers of CD4^+^ T-cells have been shown to have an improved response to BCG-therapy^13,14^. Furthermore, a study demonstrated that the urinary levels of nine different cytokines during treatment predicted the likelihood of recurrence after treatment with high accuracy^15^ indicating that multiple cell types serve a function in the immune response. No pre-treatment predictive biomarkers have been implemented for clinical use.

Upon chronic antigen stimulation from e.g. cancer or chronic viral infections, CD8 T-cells may enter a dysfunctional, exhausted state. In this state, the T-cells are characterized by sustained expression of inhibitory receptors such as PD-1, CTLA-4, and CD244 and loss of effector functions such as cytokine secretion and proliferation^16–18^. Due to the impaired immune functions of these exhausted T-cells, reversal of this state is of high therapeutic interest for improving the antitumor effects of T-cells^18^. Several immune checkpoint inhibitor therapeutics, including PD-1, PD-L1, and CTLA-4 inhibitors, target these inhibitory receptors and are now approved for treatment of various cancer types, including bladder cancer (BC)^19^. However, many tumors are either unresponsive to immunotherapies or only have a short-time benefit of the treatment, indicating the need for greater understanding of the underlying immune mechanisms^19^. Recently, the PD-1 inhibitor Pembrolizumab was approved for the treatment of BCG-unresponsive high risk bladder cancer patients with CIS based on the KEYNOTE-057 study^20^. Despite initial response rates of 41%, only ∼19% of the patients had durable responses at 12 months, highlighting the need for better understanding of the immune microenvironment.

In the current study, we integrate multiomics data, including genomics, transcriptomics, and proteomics of tumors and serial liquid biopsies from a clinically well-characterized patient cohort to delineate molecular correlates of response and resistance to BCG treatment.

## Results

### Patient characteristics and analyses

In total, 156 patients with NMIBC that received at least five instillations of BCG were retrospectively included in the study and samples were analyzed as indicated in Supplementary Fig. 2A. Patients were followed for a median of 5.5 years after BCG. Whenever possible, we included samples pre- and post-BCG. Tumors were included for WES and RNA-sequencing and urine samples for Olink and tumor derived DNA (tdDNA) analyses. Overlap in sample analyses are shown in **Supp. Fig. 2B**. See **Table 1** for clinical characteristics of patients.

**Table 1.** Clinical Characteristics - All patients

| Variable | Overall, N =<br>156<sup>1</sup> | FU < 2y, N =<br>3 | Treatment response |  |
| --- | --- | --- | --- | --- |
|  |  |  | BCG-failure, N =<br>70 | BCG-response, N =<br>83 |
| Sex, n (%) |  |  |  |  |
| Female | 33 (21%) | 1 (33%) | 17 (24%) | 15 (18%) |
| Male | 123 (79%) | 2 (67%) | 53 (76%) | 68 (82%) |
| Smoking status, n (%) |  |  |  |  |
| Current | 61 (39%) | 0 (0%) | 24 (34%) | 37 (45%) |
| Former | 74 (47%) | 2 (67%) | 36 (51%) | 36 (43%) |
| Never | 20 (13%) | 1 (33%) | 9 (13%) | 10 (12%) |
| Unknown | 1 (0.6%) | 0 (0%) | 1 (1.4%) | 0 (0%) |
| Packyears, Median (IQR) | 24 (12, 40) | 8 (6, 9) | 19 (11, 40) | 28 (15, 40) |
| Non-smokers/Unknown | 24 | 1 | 11 | 12 |
| Analyzed T-stage, n (%) |  |  |  |  |
| T1 | 47 (30%) | 2 (67%) | 24 (34%) | 21 (25%) |
| Ta | 107 (69%) | 1 (33%) | 45 (64%) | 61 (73%) |
| Ta/T1 | 2 (1.3%) | 0 (0%) | 1 (1.4%) | 1 (1.2%) |
| Analyzed grade, n (%) |  |  |  |  |
| HG | 83 (53%) | 2 (67%) | 44 (63%) | 37 (45%) |
| LG | 71 (46%) | 1 (33%) | 25 (36%) | 45 (54%) |
| LG/HG | 2 (1.3%) | 0 (0%) | 1 (1.4%) | 1 (1.2%) |
| Progression to MIBC, n (%) |  |  |  |  |
| FU < 2y | 3 (1.9%) | 3 (100%) | 0 (0%) | 0 (0%) |
| NP | 120 (77%) | 0 (0%) | 37 (53%) | 83 (100%) |
| P | 33 (21%) | 0 (0%) | 33 (47%) | 0 (0%) |
| Post-BCG follow-up (years), Median (IQR) | 5.5 (2.8, 8.2) | 1.1 (0.9, 1.4) | 4.8 (2.2, 8.4) | 6.1 (3.6, 8.1) |

<sup>1</sup> n (%); Median (IQR)
Clinical characteristics of the selected patient cohort. FU = follow-up. FU<2Y = less than two years follow-up after BCG in patients who died from other causes than bladder cancer. HG = high grade. LG = low grade+G2. NP = No progression. P = Progression

### Urinary protein profile in BCG treated patients

We analyzed 190 urine samples collected within four months before and after BCG from 124 patients, including paired samples from 66 patients. Using the Olink technique, the presence of 92 immuno-oncology related proteins was determined. We observed an upregulation of a multitude of proteins, including CXCL9, CXCL11, CXCL10, PD1, and TRAIL upon BCG treatment (**Fig. 1A; Supp. Table S1**). When stratifying the analysis according to clinical response, the general immune system activation after treatment persisted in both groups, indicating a general immune activation with BCG (higher expression of the cytokines CXCL9, CXCL10, and CXCL11; **Fig. 1B-C**). Additionally, BCG-unresponsive patients had significantly higher post-BCG levels of CD70, which has been linked to inhibition of inflammatory T-cell responses^21–23^, as well as other proteins related to immune inhibitory pathways, including PD-1, PD-L1, and CD5 (**Fig. 1C**). This was not observed in responsive patients (**Fig. 1B**). When grouping the proteins by their biological function, we observed that clinically unresponsive patients had significantly higher post-BCG levels of proteins related to chemotaxis (*p=*0.020) and showed a trend towards higher levels of proteins related to suppression of tumor immunity (*p*=0.072) and vascular and tissue remodeling pathways (*p*=0.054). No significant differences between the clinically responsive and unresponsive patients in pre-BCG samples were observed (**Fig. 1D**).

**Fig. 1.**
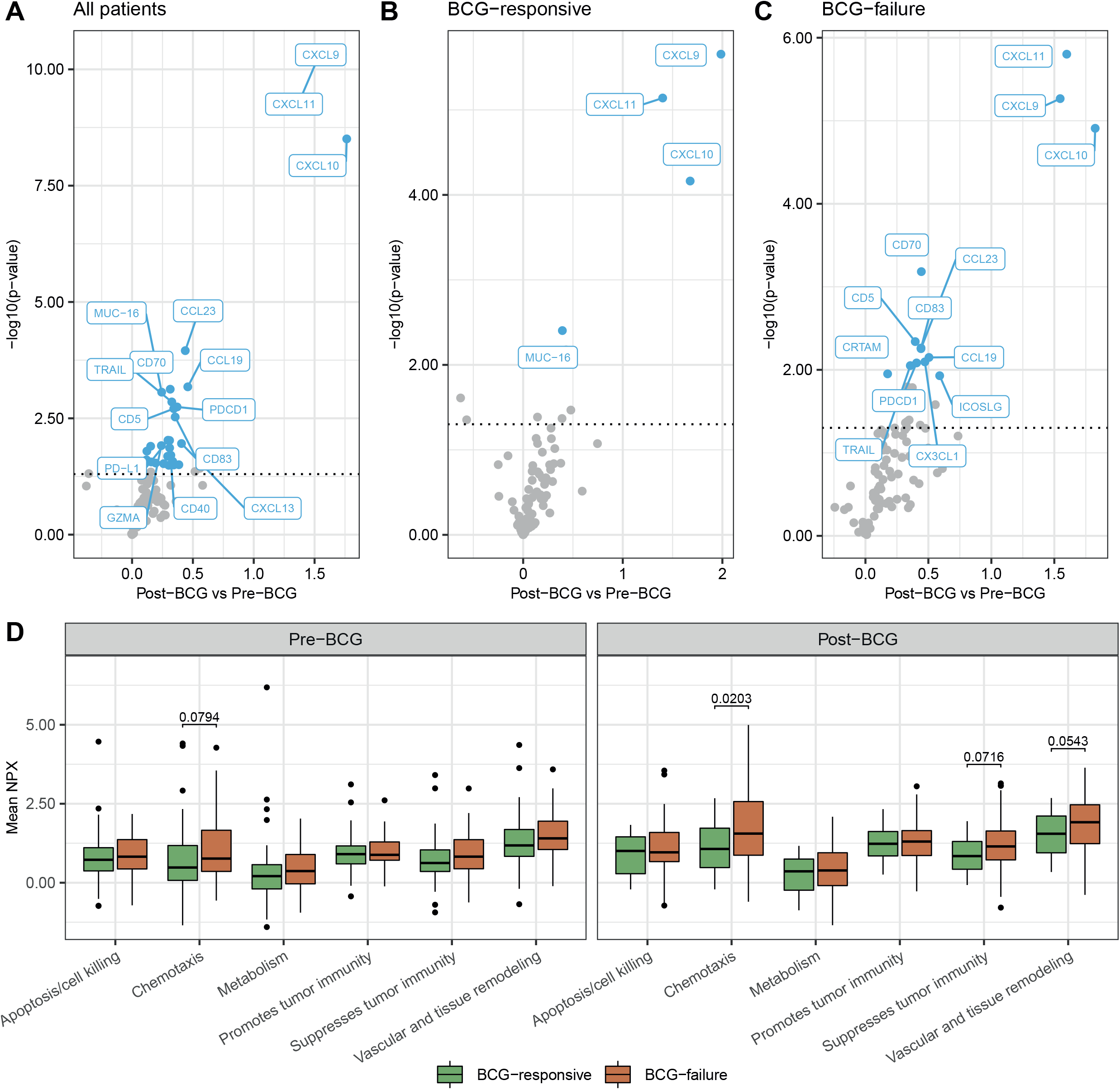
Detection of immune-oncology related responses by urine analyses. A+B+C: All or the 15 most significantly different proteins are named (Paired t-test). Blue = significant after Benjamini-Hochberg adjustment with a BH adjusted p-value (q-value) < 0.1. Grey = not significant. Dotted lines indicate a significance level of 0.05. A) Comparison between paired pre- and post-BCG urine samples (n_patients_=66). B) Comparison between pre- and post-BCG samples from BCG-responsive patients (n_patients_=27). C) Comparison between pre- and post-BCG samples from patients with BCG-failure (n_patients_=39). D) Comparison of the mean urinary level of proteins grouped by biological features from BCG-responsive and unresponsive patients in pre- and post-BCG samples (n=107 and n=83, respectively; Wilcoxon rank sum test). Non-adjusted *p*-values below 0.1 are indicated.

### Gene expression subtypes and deconvolution of immune cell populations in pre- and post-BCG tumors

A total of 170 samples (126 pre-BCG and 44 post-BCG) were classified according to the UROMOL2021 systems^24^, including 39 paired samples. The majority of pre-BCG tumors were of the high risk class 2a subtype, whereas the immune infiltrated subtype, class 2b, was relatively more common in post-BCG tumors (**Fig. 2A**). Patients had significantly different post-BCG high-grade recurrence free survival (HGRFS) depending on the UROMOL2021 pre-BCG subtypes being worse in patients with class 2a and 2b tumors (*p=*0.015; **Fig. 2B**). Furthermore, UROMOL2021 subtypes were significantly associated with clinical response to BCG (*p*=0.033) analyzed in 123 pre-BCG samples, possibly explained by a better prognosis for class 1 and class 3 tumors (**Fig. 2C**). Paired pre- and post-BCG tumors were analyzed and 43.6% of the patients remained or shifted to class 2b after BCG-treatment (**Fig. 2D**). Importantly, classifications after BCG were not associated with the time from treatment to tumor sampling (*p*=0.29, **Supp. Fig. 2A**).

We estimated immune cell populations from the RNA-seq data using established gene expression signatures^25,26^. Unresponsive patients had a higher total immune infiltration score after BCG (*p=*0.046; **Fig. 2E**). Higher post-BCG levels of dendritic cells (*p*=0.036), exhausted CD8 T-cells (*p*=0.017), and mast cells (*p*=0.043) were associated with BCG-failure, while no immune cell populations were associated with response before BCG treatment. We analyzed the expression of genes involved in selected biological processes associated with immune response and found no significantly differentially expressed genes between responsive and unresponsive patients in pre-BCG samples (**Fig. 2F**). However, after BCG, clinically unresponsive patients had significantly higher expression levels of the immune inhibitory marker genes *PD1, KLRG1, HAVCR2, CTLA4*, and *LAG3* amongst others (**Fig. 2F-G**^16^).

**Fig. 2.**
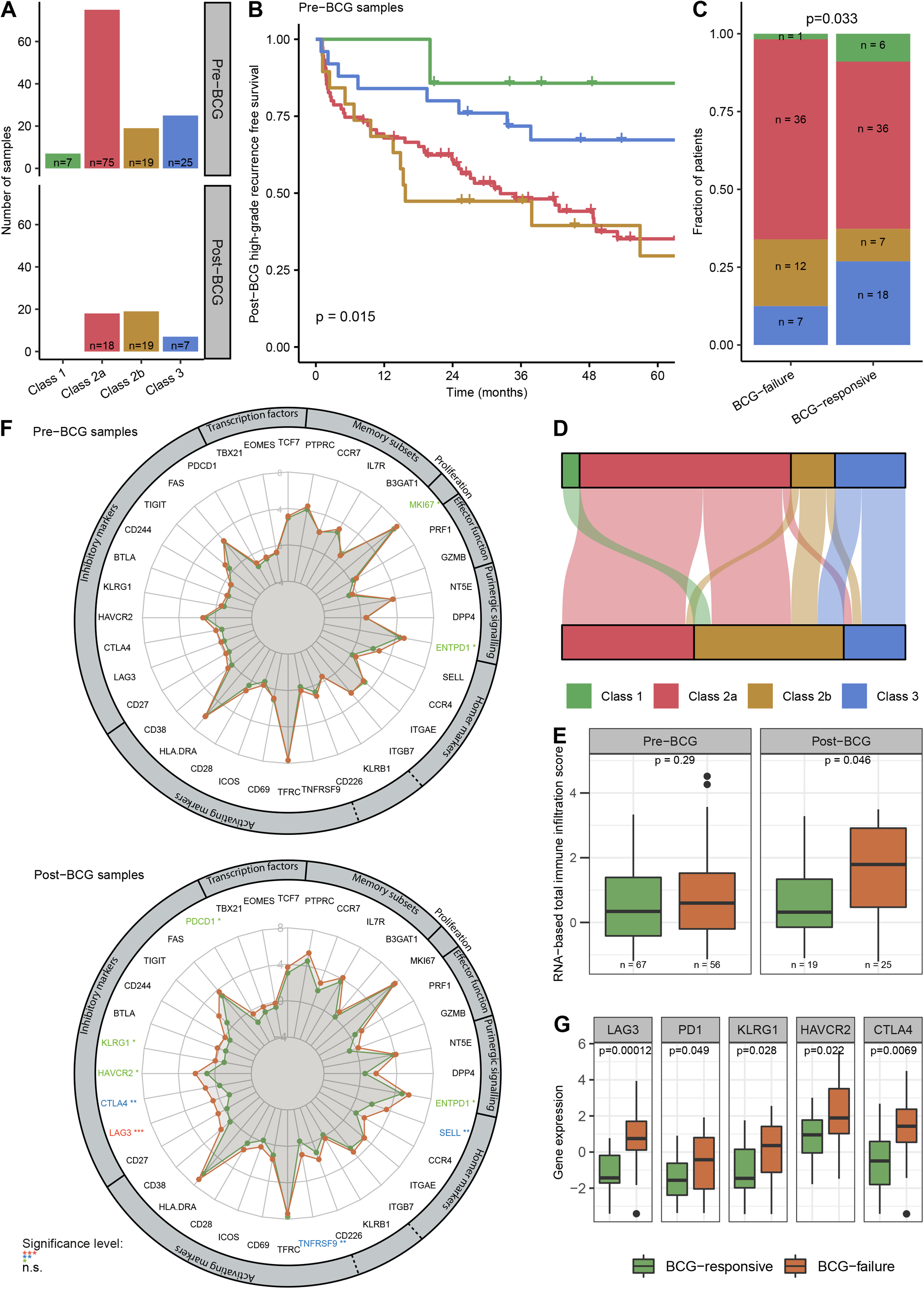
Gene expression in tumors from BCG-responsive and unresponsive patients. A) Distribution of transcriptomic classes in pre- and post-BCG tumor samples. B) Kaplan-Meier plot of post-BCG high grade recurrence free survival for 126 patients stratified by transcriptomic class in pre-BCG samples (log-rank test). C) Distribution of pre-BCG transcriptomic classes in BCG-failure and BCG-responsive patients, respectively (Fisher’s Exact Test). Three patients omitted from analysis due to lack of post-BCG FU. D) Sankey plot of transcriptomic classes before and after BCG for 39 patients. E) RNA-based immune cell infiltration in pre- and post-BCG samples from BCG-responsive and BCG-failure patients (Wilcoxon Rank Sum test). F) Spiderplot with expression level of selected immune related genes from pre-BCG samples (top) and post-BCG samples (bottom). Genes with unadjusted significantly different expression between BCG-responsive and unresponsive patients are marked by asterisks depending on significance level (Wilcoxon Rank Sum test). G) Comparison between the level of expression of selected immune inhibitory markers in BCG-responsive and BCG-failure patients post-BCG (Wilcoxon Rank Sum test).

### Exhaustion of T-cells in the tumor microenvironment

To explore the impact of CD8 T-cell exhaustion on BCG response further, we estimated an exhaustion score adjusted for CD8 T-cell infiltration for all tumors (see Methods; **Fig. 3A**). A higher level of CD8 T-cell exhaustion was associated with lack of response to BCG treatment in the post-treatment setting (pre-BCG, p=0.056; post-BCG, p=0.0021; **Fig. 3B**). Notably, post-BCG tumors with excessive CD8 T-cell exhaustion (residuals>1.05, 90th percentile) were all associated with BCG failure (*p*=0.029; **Supp. Fig. 2C-D**).

**Fig. 3.**
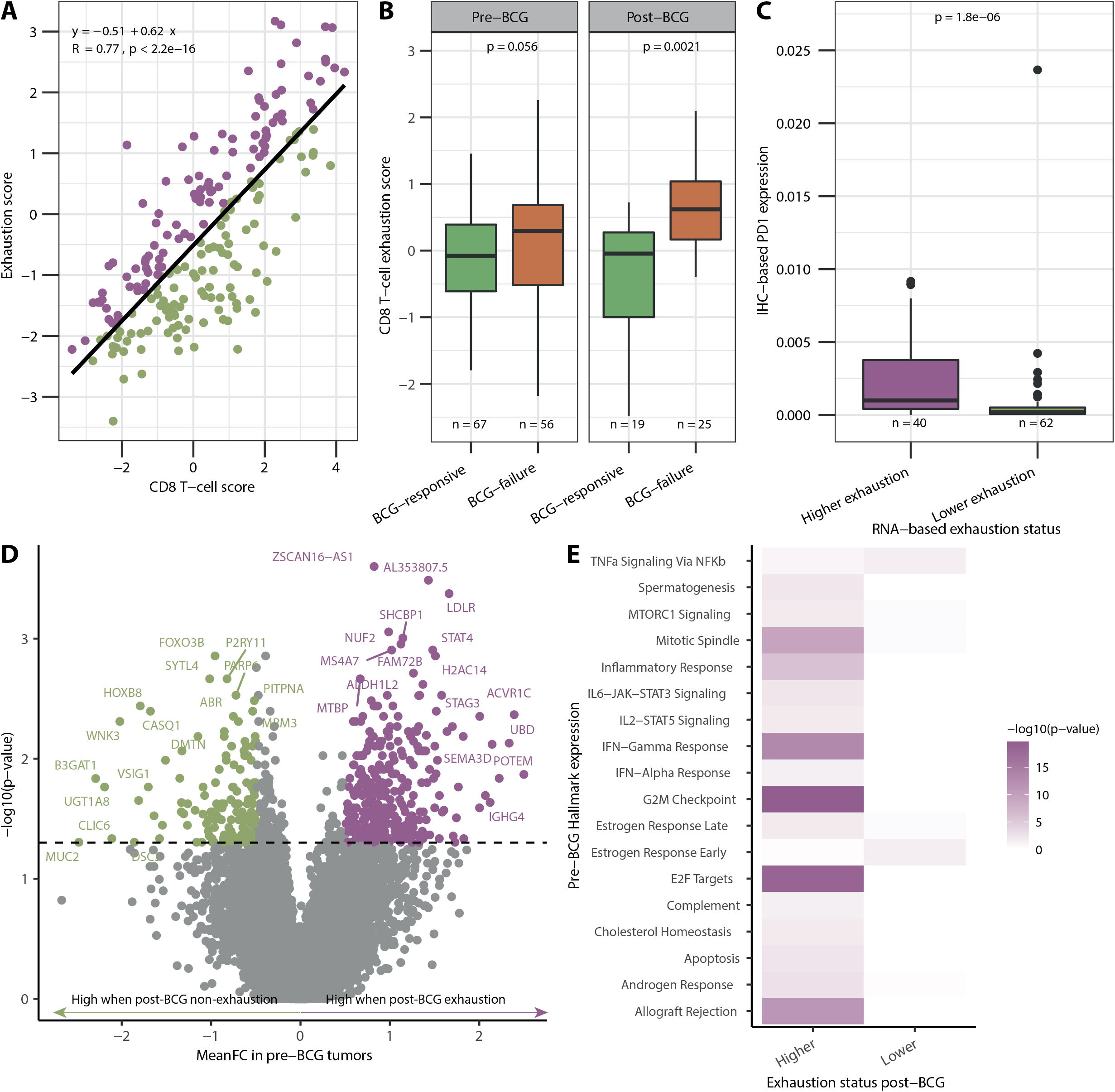
T-cell exhaustion. A) Correlation between CD8 T-cell score and exhaustion score. Green = lower exhaustion (residuals <=0.1). Purple = higher exhaustion (residuals >0.1). Pearson correlation was used to determine the correlation coefficient *R* and *p*-value. B) CD8 adjusted exhaustion score in BCG-unresponsive and responsive patients pre- and post-BCG, respectively (Wilcoxon Rank Sum test). C) Association between RNA-based exhaustion status and PD1 expression from immunohistochemistry (IHC) (Wilcoxon Rank Sum test). D) Differentially expressed genes in pre-BCG tumors from patients with higher and lower exhaustion of post-BCG tumors (n=39) identified using Wilcoxon Rank Sum test. Only genes with a mean expression>0 across all samples were included. Purple coloring indicates *p*<0.05 and mean fold change (FC)>0.5 and green coloring indicates *p*<0.05 and mean FC <-0.5. E) Hallmark pathways significantly upregulated in pre-BCG tumors from patients with higher or lower exhaustion of post-BCG tumors, respectively (Fisher’s exact test).

To validate the CD8 T-cell exhaustion score, we applied the same approach on the UROMOL2021 cohort. These samples were also analyzed by multiplex immunofluorescence and immunohistochemistry to determine the level of immune cell infiltration and PD1/PD-L1 protein expression. We observed that high PD1 protein expression was significantly associated with higher RNA-based exhaustion score (*p*=1.8×10^−6^; **Fig. 3C**).

To identify possible pre-BCG markers of post-BCG CD8 T-cell exhaustion, we identified differentially expressed genes between pre-BCG tumors from patients with higher and lower exhaustion in post-BCG tumors (**Fig. 3D**). Using the significant genes with a mean fold change >0.5, we found that especially cell cycle- and immune response-related pathways were significantly enriched in pre-BCG tumors from patients with higher exhaustion in post-BCG tumors, including G2M checkpoint (p=2×10^−20^), E2F targets (p=3×10^−19^), interferon gamma response (p=7×10^−14^), and inflammatory response (p=1×10^−6^; **Fig. 3E**).

### Genomic correlates to response and exhaustion

We sought to identify genomic features of pre-BCG tumors predictive of treatment response and exhaustion. Pre-BCG tumors from BCG-responsive patients demonstrated fewer INDELs compared to tumors from unresponsive patients (*p*=0.045; **Supp. Fig. S3**). No differences were observed for single nucleotide variants (SNVs; *p*=0.77). We performed *de novo* extraction of mutational signatures and identified six, but no association with response to treatment or a high level of CD8 T-cell exhaustion after BCG was observed (**Supp. Fig. S3-5**). No other genomic features in pre-BCG tumors were associated with a high level of CD8 T-cell exhaustion after BCG (**Supp. Fig. S4**). Furthermore, we performed a gene level permutation test to assess if mutation status of specific genes in pre-BCG samples was predictive of post-BCG CD8 T-cell exhaustion and found, amongst others, *MUC4* to be more frequently mutated in the post-BCG high-exhaustion group (*p*=0.0007) and *FGFR3* in the low-exhaustion group (*p*=0.056; **Fig. 4A**). Furthermore, we investigated correlations between the level of neoantigens in the tumors and exhaustion status. Patients with a high neoantigen load in pre-BCG tumors had significantly higher levels of exhaustion in post-BCG tumors (*p*=0.023; **Fig. 4B**).

**Fig. 4.**
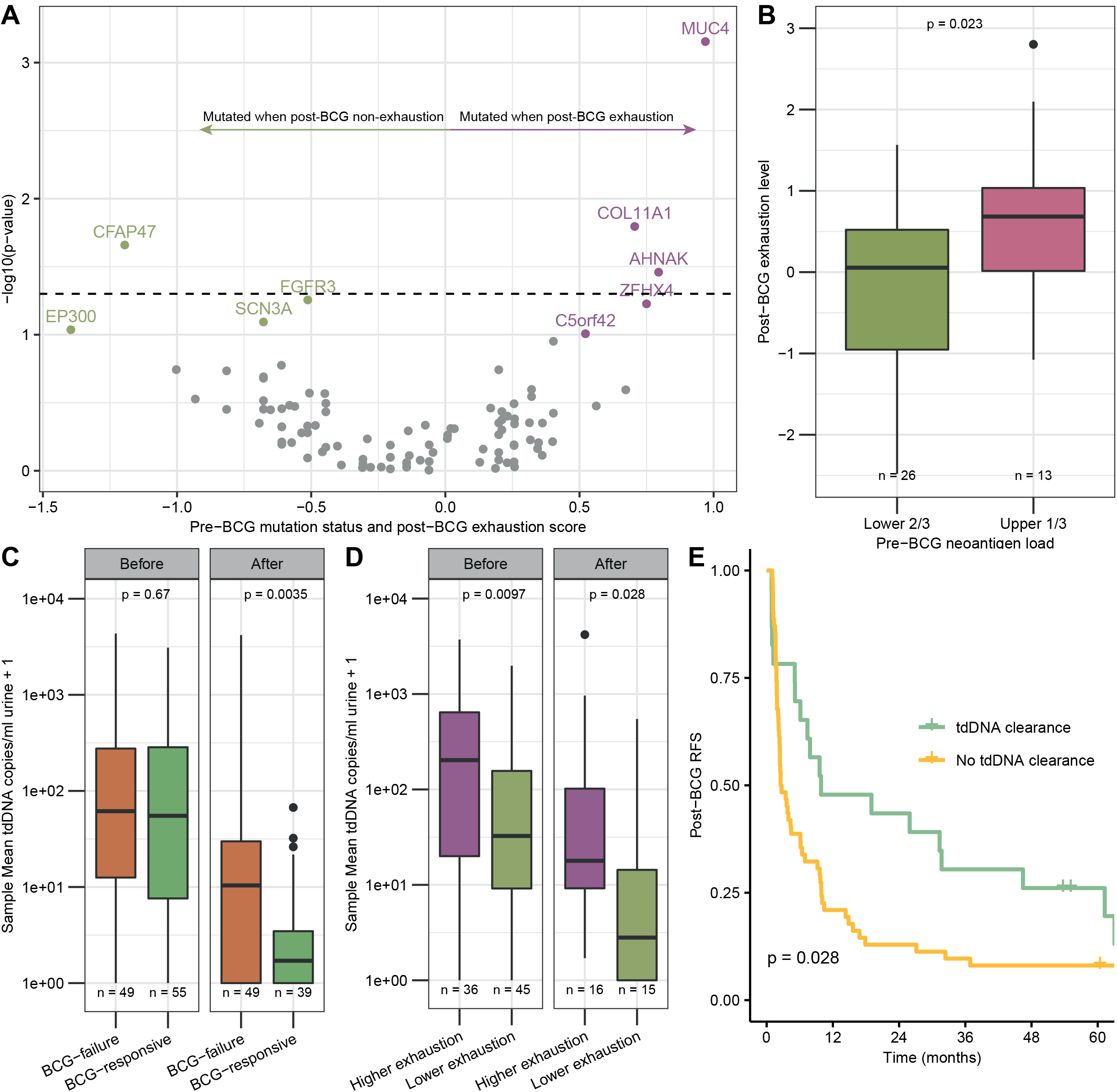
Genomics and BCG response. A) Gene level permutation test on mutation status of specific genes and post-BCG CD8 T-cell exhaustion (Wilcoxon Rank Sum test). B) Neoantigen load (upper 1/3 vs lower ⅔) in pre-BCG tumors correlated to exhaustion level in post-BCG tumors (Wilcoxon Rank Sum test). C) A per sample mean of urinary tdDNA copies per ml (one is added to all sample levels) in all pre- and post-BCG urine samples from BCG responsive and unresponsive patients (Wilcoxon Rank Sum test). D) A per sample mean of urinary tdDNA copies per ml (one is added to all sample levels) in all pre- and post-BCG urine samples and exhaustion status of pre- and post-BCG tumors, respectively (Wilcoxon Rank Sum test). E) Kaplan-Meier plot of post-BCG RFS for patients with and without tdDNA clearance post-BCG (log-rank test).

### Urinary tdDNA as a measure of response

To investigate possible correlations between response to BCG and urinary tdDNA levels and dynamics, we analyzed 192 pre- and post-BCG urine samples from 107 patients, including 85 paired samples, by deep-targeted sequencing of tumor-specific mutations. When analyzing all pre-BCG and post-BCG samples, we observed significantly higher post-BCG levels of tdDNA in unresponsive patients (*p*=0.0035), while there was no difference in pre-BCG samples (*p*=0.67; **Fig. 4C**). The exhaustion status of pre- and post-BCG tumors was significantly associated with the levels of tdDNA both in pre- and post-BCG urine samples (*p*=0.0097 and *p*=0.028, respectively; **Fig. 4D**). Furthermore, when focusing on paired samples, we observed that patients with tdDNA clearance after BCG had significantly better post-BCG RFS compared to patients without tdDNA clearance (*p*=0.028; **Fig. 4E**).

### Pre-BCG predictors of post-BCG exhaustion and BCG-failure

To further examine the biological differences between BCG-responsive and BCG-unresponsive patients, we analyzed the prognostic and predictive role of identified pre-BCG predictors of post-BCG CD8 T-cell exhaustion and BCG-response as well as of clinical variables. Based on the ratio between differentially expressed genes in pre-BCG tumors, a post-BCG exhaustion predictor (post-BCG ExhP) was established. Univariate Cox Regression analysis showed that a high post-BCG ExhP (*p*=0.0084), UROMOL class 2a and 2b (*p*=0.0023), and the pre-BCG CD8 T-cell exhaustion score (*p*=0.047) were associated with a worse post-BCG HGRFS, possibly indicating BCG-failure, whereas *FGFR3* mutations were associated with better post-BCG HGRFS (*p*=0.011; **Fig. 5A**). No clinical variables were associated with post-BCG HGRFS. A simplified schematic representation of identified pre-BCG predictors of response and post-BCG exhaustion is illustrated in **Fig. 5B**.

**Fig. 5.**
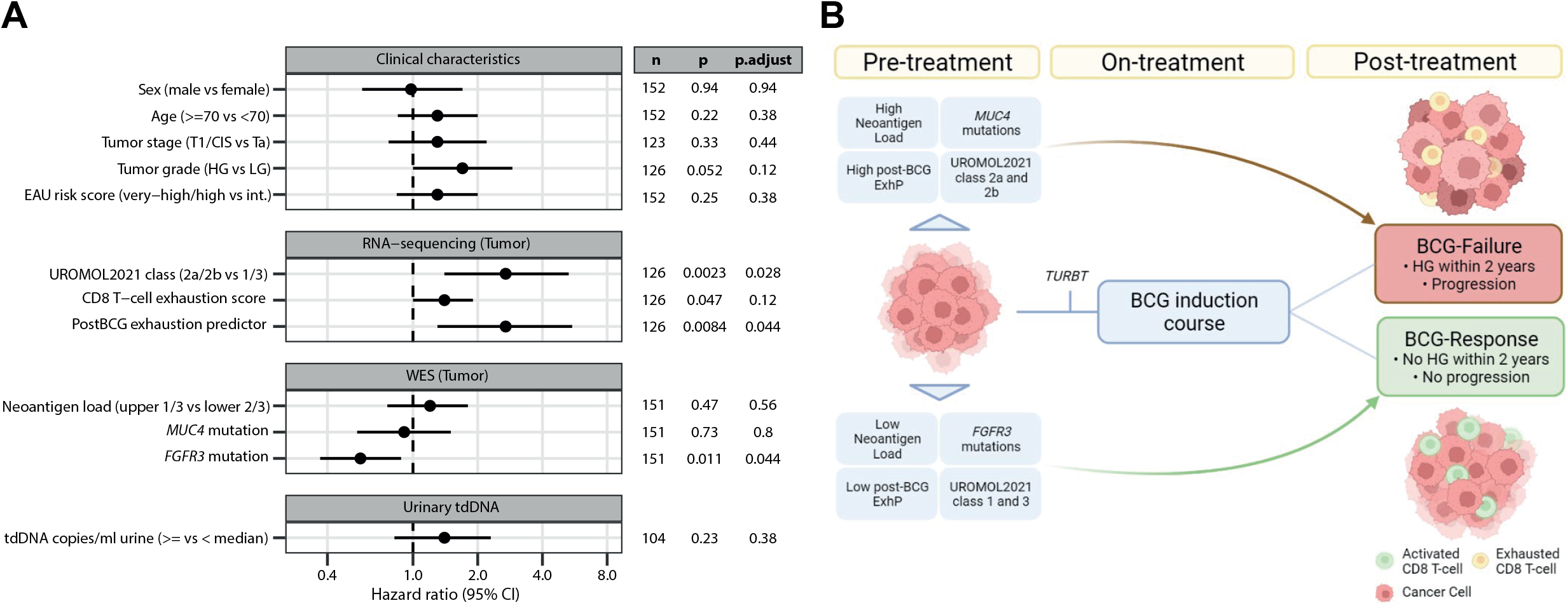
Transcriptomic and genomic features predict post-BCG HGRFS. A) Forest plot of hazard ratios calculated from univariate Cox regressions of post-BCG HGRFS using clinical and molecular features of pre-BCG tumors or pre-BCG urine characteristics. Dots represent hazard ratios and horizontal lines the corresponding 95% confidence intervals (95% CI). *P*-values are calculated by the Wald test. Benjamini Hochberg correction was applied for adjustment of multiple testing (p.adjust). n indicates the number of patients in each analysis. EAU risk was calculated for the BCG-inducing tumor. B) Simplified schematic representation of identified pre-BCG predictive variables of T-cell exhaustion and BCG-failure. LG = Low grade. HG = high grade. TURBT = Transurethral resection of bladder tumor. Created with BioRender.com (publication license obtained).

## Discussion

Here we performed multiomics analysis and identified T-cell exhaustion as a possible explanation for BCG-unresponsiveness observed in both urine and tumor samples. We established a CD8 T-cell adjusted exhaustion score and demonstrated that patients with BCG-failure had significantly higher exhaustion levels. Following, we developed a post-BCG exhaustion prediction (post-BCG ExP) score from pre-BCG gene expression levels related to post-BCG exhaustion. We showed that high post-BCG ExP scores were predictors of worse post-BCG HGRFS, indicating BCG-failure. Furthermore, pre-BCG tumors of patients with post-BCG exhaustion had higher expression of genes in cell cycle and immune related pathways, possibly indicating a more aggressive and immune active phenotype. More actively dividing cancer cells potentially leads to increased mutagenesis. Through generation of neoantigens, mutations have been shown to alter the tumor immunogenicity by increasing T-cell responses^27^. However, chronic stimulation of T-cells, e.g. by neoantigens, increases the risk of T-cells entering the dysfunctional state of exhaustion^17^, indicating a delicate balance between T-cell activation resulting in tumor elimination and T-cell overstimulation leading to dysfunction and exhaustion. The importance of distinguishing between “inflamed” versus “noninflamed” and antitumor-versus protumerigenic inflammation, a “Hallmark of Cancer”^28^, has been suggested by Wang *et al*.^*29*^, highlighting the complexity of the fine balances in the TME. Our results support this immunological balance and show that BCG treatment in some patients leads to exhaustion and BCG-unresponsiveness, which has also been suggested by others^30^. Furthermore, we observed that high neoantigen loads in pre-BCG tumors were correlated to higher levels of post-BCG exhaustion. Hence, the risk of tipping the balance towards a protumerigenic TME may be more pronounced in patients where pre-treatment tumors are characterized by high activity of cell division and immunological pathways and with high neoantigen loads. In Lindskrog *et al*.^*24*^ it was hypothesized that class 2b tumors respond poorly to BCG due to high expression of immune checkpoint markers and should therefore be offered treatment with immune checkpoint inhibitors. In the current study, we found pre-BCG class 2a and 2b tumors to have the worst post-BCG HGRFS, compared to class 1 and class 3 tumors, which may simply reflect the prognostic value of the classification system. Though not significant (only 19 class 2b tumors vs 75 class 2a), we did observe a separation of the two classes within two years after BCG with class 2b having the worst post-BCG HGRFS. Based on our current study we cannot determine if Class 2b has a worse prognosis after BCG, and additional larger studies are needed.

*MUC4* was mutated significantly more often in pre-BCG tumors from patients with post-BCG exhaustion. In a recent study on colon cancer, mutations in *MUC4* were linked to activation of immune system signaling pathways and enhancement of the antitumor immune response^31^. This link might explain the observed correlation to exhaustion through BCG-induced overactivation of the already active immune system. To the contrary, *FGFR3* mutations have been linked to non-inflamed tumors suggesting a role of FGFR3 in T-cell exclusion in BC^32,33^. This may result in an exhaustion-protective role of *FGFR3* mutations explaining the correlation to low post-BCG exhaustion.

Circulating tumor DNA (ctDNA) has shown promising results in predicting relapse after treatment in BC and for monitoring response during treatment in MIBC^34–36^. Urinary tdDNA may reflect both local bladder neoplastic disease as well as disseminated disease where tdDNA is present in the urine after renal clearance^37^ making monitoring of tdDNA potentially useful in a NMIBC setting. In the current study, we observed that the levels and dynamics of tdDNA were associated with disease outcome after BCG treatment. Presence of tdDNA after treatment, even in the absence of visible tumor, identified patients at high risk of recurrence, suggesting that these patients might benefit from intensified treatment. The low levels of tdDNA after BCG can be a result of both surgical removal of the tumor and the effect of BCG treatment. However, since all patients received both treatments and differences between BCG-responsive and -unresponsive patients were observed, tdDNA may have a predictive role that needs to be investigated further.

Validation of the presented findings are necessary to establish the clinical significance. To distinguish prognostic and predictive markers and establish the true predictive power, a comparison study is necessary. Biobank data, as presented in this study, are not suitable for this since all patients have received BCG. However, a future randomized trial including the markers would clarify this.

Altogether, these results could be used to guide clinical decision making, if validated in clinical trials. A possible intervention could be treatment with immune-modulatory agents in combination with BCG in BCG-naïve or unresponsive patients as is currently being investigated in clinical trials^38,39^. Furthermore, close follow-up of patients by consecutive liquid biopsy monitoring during and after treatment to continue, escalate or deescalate treatment upon indications of exhaustion of the T-cells or lack of response to treatment is a clinically applicable method, where the advantages of liquid biopsies can be fully used.

## Conclusions

Our results highlight several correlates to BCG-failure, and importantly we show that CD8 T-cell exhaustion may be a key factor of this. Pre-BCG prediction of post-BCG CD8 T-cell exhaustion has the potential to identify patients at high risk of BCG-failure and urinary measurements of proteins and tdDNA to monitor treatment response. If validated in clinical trials, this could potentially improve treatment regimens for patients with high-risk NMIBC.

## Supporting information

Supplementary figures, tables as well as materials and methods

## Data Availability

Sharing of sensitive pseudonymized patient specific clinical information and any raw sequencing data is currently not possible due strict ethics and GDPR regulations.

## Author contributions

Lars Dyrskjøt had full access to all the data in the study and takes responsibility for the integrity of the data and the accuracy of the data analysis.

### Study concept and design

Strandgaard, Dyrskjøt.

### Acquisition of data

Strandgaard, Nordentoft, Andreasen, Ranti, Wang, Bieber, Sfakianos, Horowitz, Jensen.

### Analysis and interpretation of data

Strandgaard, Lindskrog, Nordentoft, Christensen, Birkenkamp-Demtröder, Lamy, Kjær, Jensen, Dyrskjøt.

### Drafting of the manuscript

Strandgaard, Lindskrog, Dyrskjøt.

### Critical revision of the manuscript for important intellectual content

All authors

### Statistical analysis

Strandgaard, Lindskrog, Christensen, Kjær.

### Obtaining funding

Strandgaard, Dyrskjøt.

### Administrative, technical, or material support

Andreasen, Rasmussen, Steiniche, Jensen, Dyrskjøt.

### Supervision

Dyrskjøt.

### Other

None.

## Financial disclosures

Lars Dyrskjøt has sponsored research agreements with C2i, AstraZeneca, Natera, Photocure, and Ferring; has an advisory/consulting role at Ferring; and is Chairman of the Board in BioXpedia A/S. Jørgen Bjerggaard Jensen is proctor for Intuitive Surgery; is a member of advisory board for Olympus Europe, Cephaid, and Ferring; and has sponsored research agreements with Medac, Photocure ASA, Cephaid, and Ferring.

## Funding/Support and role of the sponsor

This work was funded by Ferring Pharmaceuticals, the Danish Cancer Society, Aarhus University, Dagmar Marshalls Fond, Christian Larsen og Dommer Ellen Larsens Legat, Fabrikant Einar Willumsens Mindelegat, the Danish Medical Association, Danish Cancer Biobank, and Dansk Kræftforskningsfond.

## Acknowledgements

We thank all technical personnel at the Department of Molecular Medicine, Aarhus University Hospital, for sample handling and processing.

## Materials and methods

See Supplementary Materials and Methods for further descriptions.

### Patients, biological samples, and clinical follow-up

All patients were treated at Aarhus University Hospital, Denmark, between 1996 and 2021 and provided informed written consent to take part in future research projects. The study was approved by The Danish National Committees on Health Research Ethics (#1708266). A total of 156 patients with NMIBC were retrospectively selected for the study. Analyses were focused around the first induction course consisting of at least five instillations of BCG. Ten patients received at least three instillations of BCG-maintenance treatment following the induction course. In this study with few patients treated with maintenance-BCG, “BCG-failure” was defined as recurrence of high grade (HG) urothelial carcinomas within two years after end of BCG induction treatment or progression to muscle-invasive bladder cancer (MIBC) any time during follow-up (FU). Patients who died from other causes than BC less than two years after BCG and who were otherwise defined as BCG-responsive, were omitted from BCG-response and progression analyses (n=3).

Samples were collected before and after BCG. A total of 198 tumors were analyzed using WES (151 pre-BCG, 47 post-BCG), 170 tumors using RNA sequencing (126 pre-BCG, 44 post-BCG), 190 urine supernatants with Olink proteomics (107 pre-BCG, 83 post-BCG), and 192 urine supernatants (104 pre-BCG, 88 post-BCG) with deep-targeted sequencing. All analyses utilizing clinical variables were performed with the available sample closest to BCG initiation and completion for each patient (**Supp. Fig. 1**).

End of FU was defined as the last of the following: last cystoscopy, last detected tumor, cystectomy, progression or metastases. Progression-free-survival (PFS) and recurrence-free-survival (RFS) were calculated from analyzed pre-BCG tumor closest to BCG-start to recurrence or progression, respectively, or end of FU. Post-BCG RFS and High-grade recurrence free survival (HGRFS) were calculated from BCG end-date and until first recurrence or high grade recurrence, respectively, or end of FU.

Tumor DNA was extracted from serial cryosections of fresh-frozen (FF) or punches of formalin-fixed paraffin-embedded (FFPE) tumors with Gentra Puregene Tissue Kit (Qiagen) or AllPrep DNA/RNA Kit (Qiagen), respectively. Leukocyte DNA for germline (GL) reference was extracted using Qiasymphony DSP DNA midi kit (Qiagen). Total Tumor RNA was extracted from serial cryosections using RNeasy Mini and Micro Kits (Qiagen). Urine supernatant samples were processed and cfDNA extracted as described previously using QIAsymphony DSP Circulating DNA Kit (Qiagen)^35^.

### Urine proteomic analysis

Urine samples were analyzed for the presence of 92 immuno-oncology related proteins (Supplementary Table S1) using the Olink Target 96 Immuno-Oncology Assay (Olink). In short, these measurements are based on the Proximity Extension Assay (PEA) technology which enables high-throughput multiplex immunoassays of proteins using 1 µl of urine. Normalized Protein eXpression (NPX) log-2 normalized data provided by Olink^®^ was used for data analysis.

### RNA-sequencing

RNA-seq was performed using either ScriptSeq (EpiCentre) library preparation followed by sequencing on an Illumina HiSeq 2000 or using the KAPA RNA HyperPrep Kit (RiboErase HMR; Roche) for library preparation followed by sequencing on an Illumina NovaSeq6000.

Salmon was used to quantify the expression of transcripts using annotation from the Gencode release 33 on genome assembly GRCh38. The tximport R library was then used to import the transcript-level estimates and summarize them at the gene-level. Finally, samples with less than 5 mio. mapped reads were excluded and genes not expressed in more than 25% of the remaining samples were filtered out.

### Whole Exome Sequencing

Whole Exome Sequencing (WES) library construction and capture was performed using the Illumina TruSeq DNA kit followed by SeqCap EZ v3.0 capture or by Twist Library preparation EF kit followed by Twist Human Core Exome Capture kit combined with refSeq spike-in probes from Twist Bioscience. Samples were sequenced using Illumina Sequence platforms.

Fastq files were trimmed using cutadapt and mapped with bwa-mem using the GRCh38 genome assembly. Duplicate reads were marked using MarkDuplicates from GATK and base quality scores were recalibrated (ApplyBQSR, GATK). Variants were called using Mutect2.2 and annotated using SnpEffv4.3i. Finally, variants with frequency below 5% (VAF < 5%), less than three alternate allele reads in the tumor or a ROQ score (Phred-scaled probability that the variant alleles are not due to a read orientation artifact) below 30 were filtered out.

HLA-types were called using POLYSOLVER, xHLA, and OptiType. Final call made by consensus vote between the algorithms. A mutation was considered a neoantigen if at least one novel 9-11mer had an EL-rank-percentage<2, predicted by NetMHCpan-4.1.

### Deep-targeted sequencing of urine tdDNA

For deep-targeted sequencing of tumor-specific mutations in urine supernatants, we designed three tumor-guided NGS panels with 11-71 patient-specific mutations (TWIST Bioscience).

NGS libraries were prepared using a modified version of the Mechanical Fragmentation Library Preparation Kit (Max. 100ng input; Twist Bioscience) followed by a modified Twist Target Enrichment protocol (Twist Bioscience) in combination with the custom Twist panels. Unique Molecular Identifiers (UMIs) were incorporated in library preparation. UMI consensus base calls were called with the fgbio tool package (v1.4.0) with the ClipBAM function enabled. DeepSNV was applied for detection and quantification of low-frequency tdDNA mutations in urine samples. tdDNA clearance was defined as a decrease in mean variant allele fraction (VAF) per sample to zero in post-BCG samples.

### Gene expression signatures

Tumor samples were classified according to the UROMOL2021 system^24^ using the R package classifyNMIBC. RNA-based immune cell populations were estimated using established gene expression signatures^25,26^. Gene lists are available in **Supp. Table S2**. The residuals from the linear regression between the mean expression of the five genes related to immune inhibitory processes (*PDCD1, CTLA4, LAG3, HAVCR2*, and *KLRG1*; **Fig. 2G**) and the estimated level of CD8 T-cells were used as indicators of the CD8 T-cell adjusted exhaustion level for all tumors. Tumors with residuals >0.1 were defined as having higher CD8 T-cell exhaustion and tumors with residuals >1.05 (90th percentile) were defined as having excessive CD8 T-cell exhaustion.

Differentially expressed genes with a mean expression >0 across all pre-BCG tumors from patients with exhausted or non-exhausted tumors after BCG treatment (n=39) were identified using Wilcoxon Rank Sum test. Significant genes with a mean fold change >0.5 (n=306) and <-0.5 (n=143) were marked as being upregulated in pre-BCG tumors from patients having exhausted and non-exhausted tumors post-BCG, respectively. To explore pathways enriched in each selected gene list, gene lists were compared to the HALLMARK gene signatures in the Molecular Signatures Database using Fisher’s exact test (R packages msigdbr and GeneOverlap). We defined a postBCG exhaustion predictor as the ratio between the previously identified genes upregulated in pre-BCG tumors from patients having exhausted and non-exhausted tumors post-BCG, respectively.

### Statistical analysis

Fisher’s Exact test was used for categorical variables and Wilcoxon Rank Sum test (unpaired data) or Wilcoxon Signed Rank test (paired data) were used for continuous variables. Paired t-test was used for the Olink data. Cox proportional hazard regression models were performed using the R package survival. The Benjamini Hochberg correction was applied to correct for multiple testing. Statistical significance was set at p<0.05 and q<0.1. All statistical analyses were performed using R version 4.1.1.

## Notes

### Author Declarations

The Danish National Committees on Health Research Ethics gave ethical approval for this work (#1708266).

