## Supplementary figures, tables as well as materials and methods for "Elevated T-cell exhaustion and urinary tumor DNA levels are associated with BCG failure in patients with non-muscle invasive bladder cancer"

### Supplementary Fig. S1

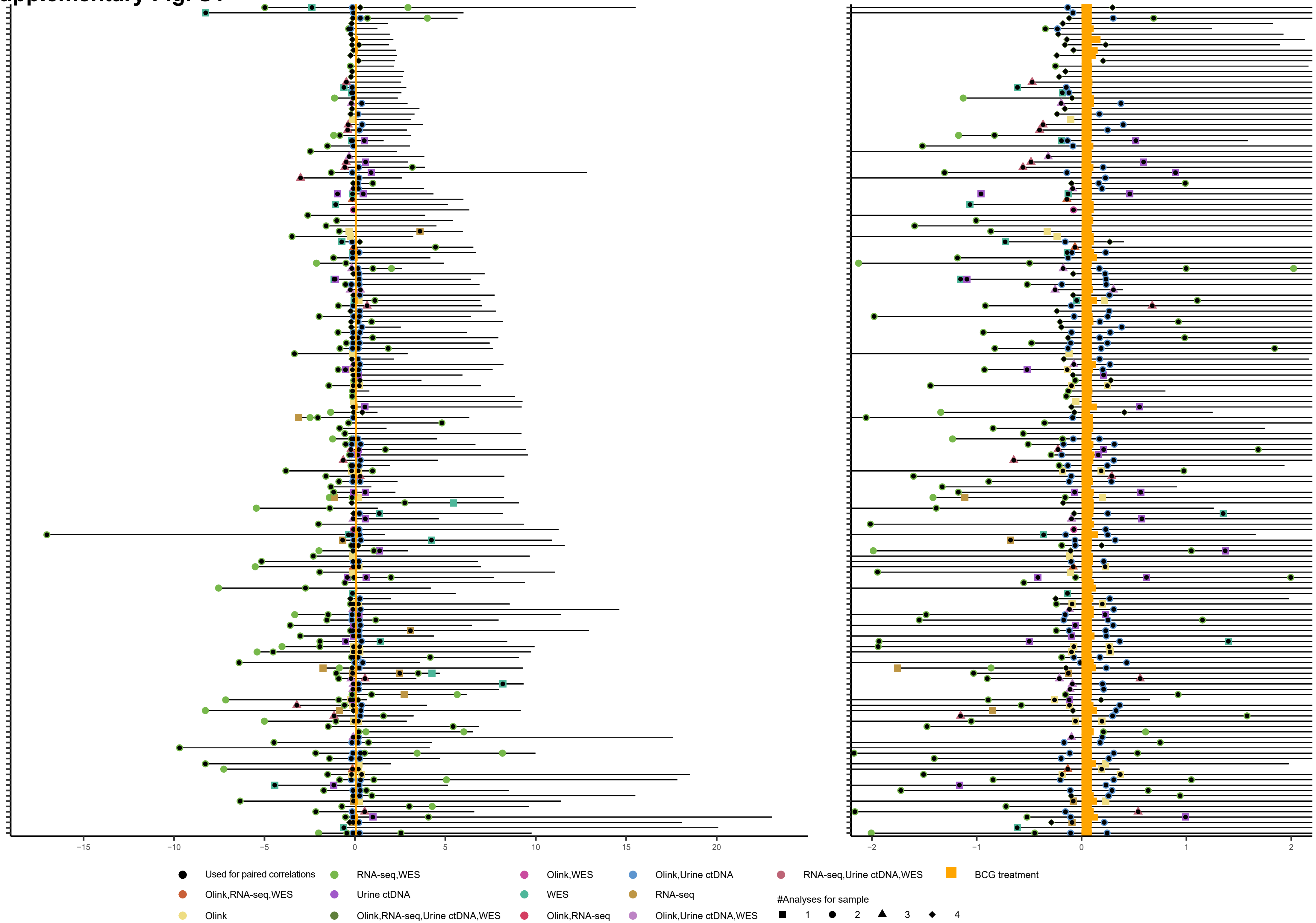

#### Supplementary Fig. S1

Detailed overview of patients disease courses and analyzed samples. Lines represent patient’s disease courses from time of first analyzed tumor or urine sample to end of FU centered around the first induction treatment of BCG with at least five instillations. Colors indicate performed analyses. Shapes indicate the number of analyses per clinical timepoint. Left: full FU. Right: Two years pre and post-BCG illustrated.

Supplementary Fig. S2

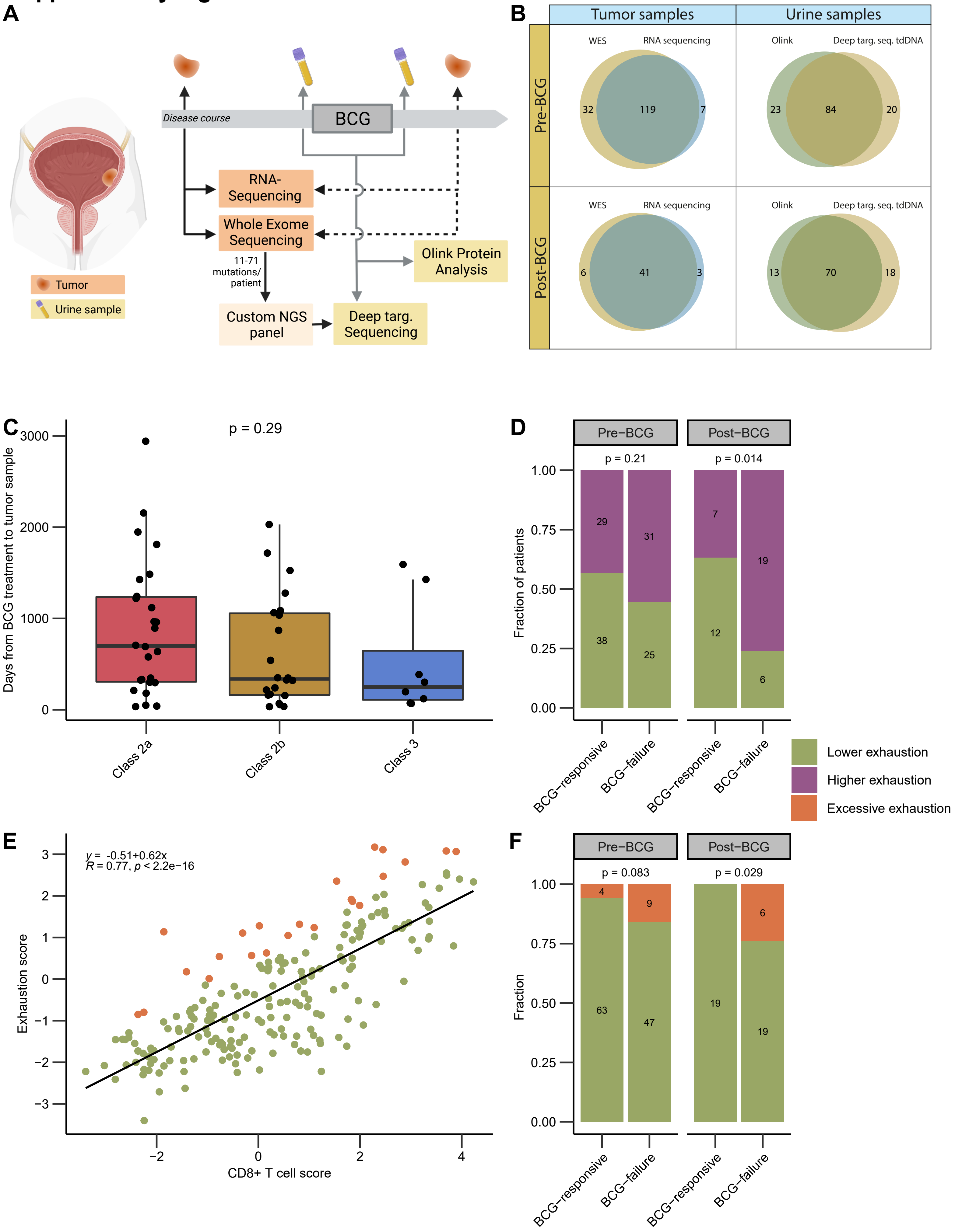

### Supplementary Fig. S2

Study and sample overview and RNAseq based analyses. A) Overview of study with disease course including BCG treatment of at least five instillations and sample types at different time points. Dashed lines indicate samples analyzed from a limited number of patients. Design of custom panel for deep targeted sequencing is outlined. Created with BioRender.com (publication license obtained). B) Venn diagram showing overlapping samples between analysis methods. C) Association between UROMOL2021 tumor subtype and time from BCG treatment for all post-BCG samples sequenced for the project (Wilcoxon Rank Sum test). D) CD8 adjusted exhaustion status in BCG-responsive and BCG-failure patients in pre- and post-BCG tumors, respectively (Fisher's Exact test). E) Correlation between CD8 T-cell score and exhaustion score. Green = lower exhaustion (residuals <=1.04). Purple = excessive exhaustion (residuals >1.04). Pearson correlation was used to determine the correlation coefficient *R* and *p*-value. F) Excessive exhaustion status in BCG-responsive and BCG-failure patients in pre- and post-BCG tumors, respectively (Fisher's Exact test).

### Supplementary Fig. S3

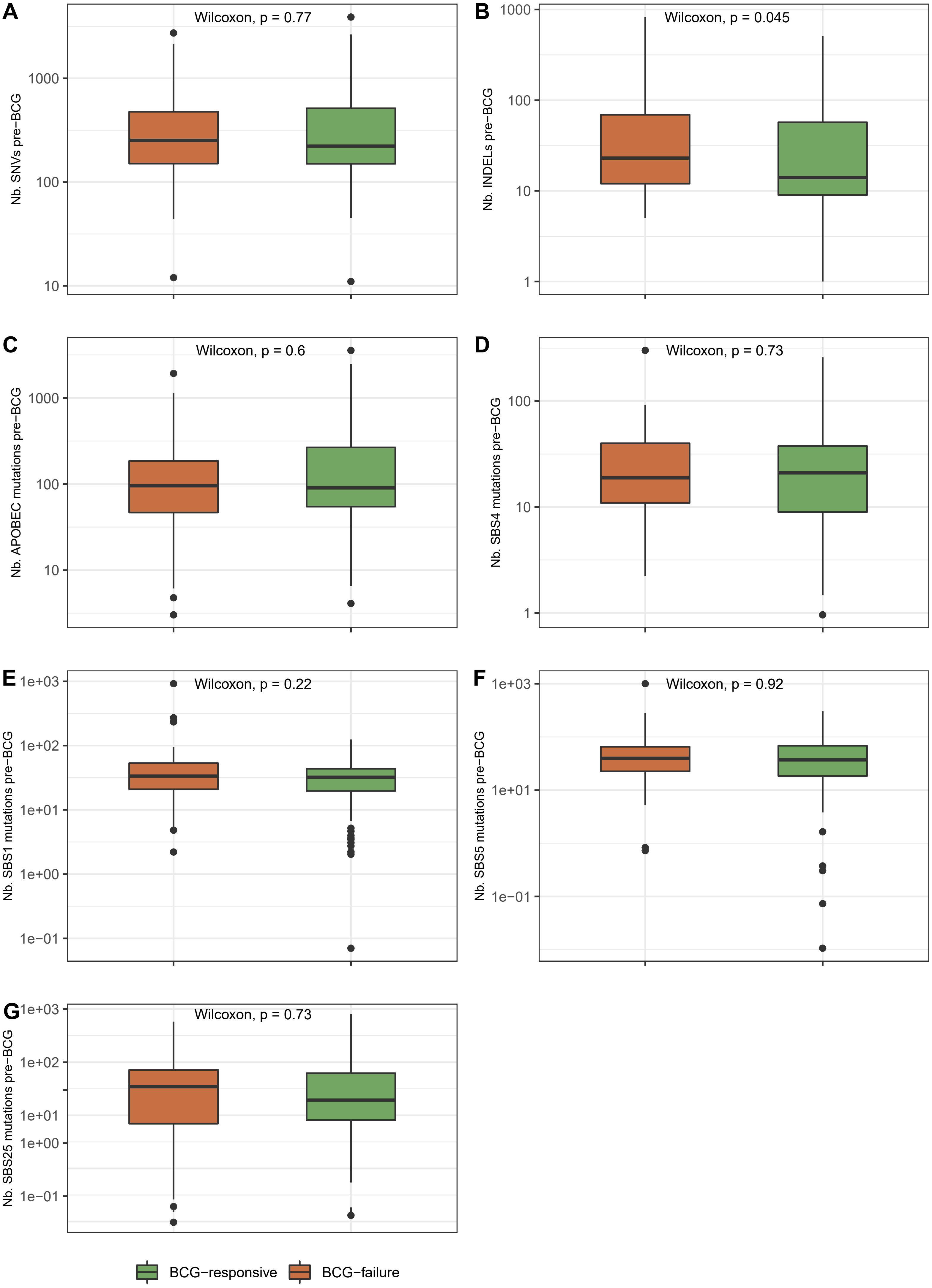

#### Supplementary Fig. S3

Correlations between tumor WES characteristics and BCG-response. A) The number of single nucleotide variants (SNVs) in pre-BCG tumor samples correlated to response to BCG. B) The number of INDELs in pre-BCG tumor samples correlated to response to BCG. C) The number of APOBEC-related mutations (SBS2+SBS13) in pre-BCG tumor samples correlated to response to BCG. D) The number of SBS4-related mutations in pre-BCG tumor samples correlated to response to BCG. E) The number of SBS1-related mutations in pre-BCG tumor samples correlated to response to BCG. F) The number of SBS5-related mutations in pre-BCG tumor samples correlated to response to BCG. G) The number of SBS25-related mutations in pre-BCG tumor samples correlated to response to BCG. Green = BCG-responsive patients. Orange = BCG-failure patients. The Wilcoxon Rank Sum test was used for statistical comparisons.

#### Supplementary Fig. S4

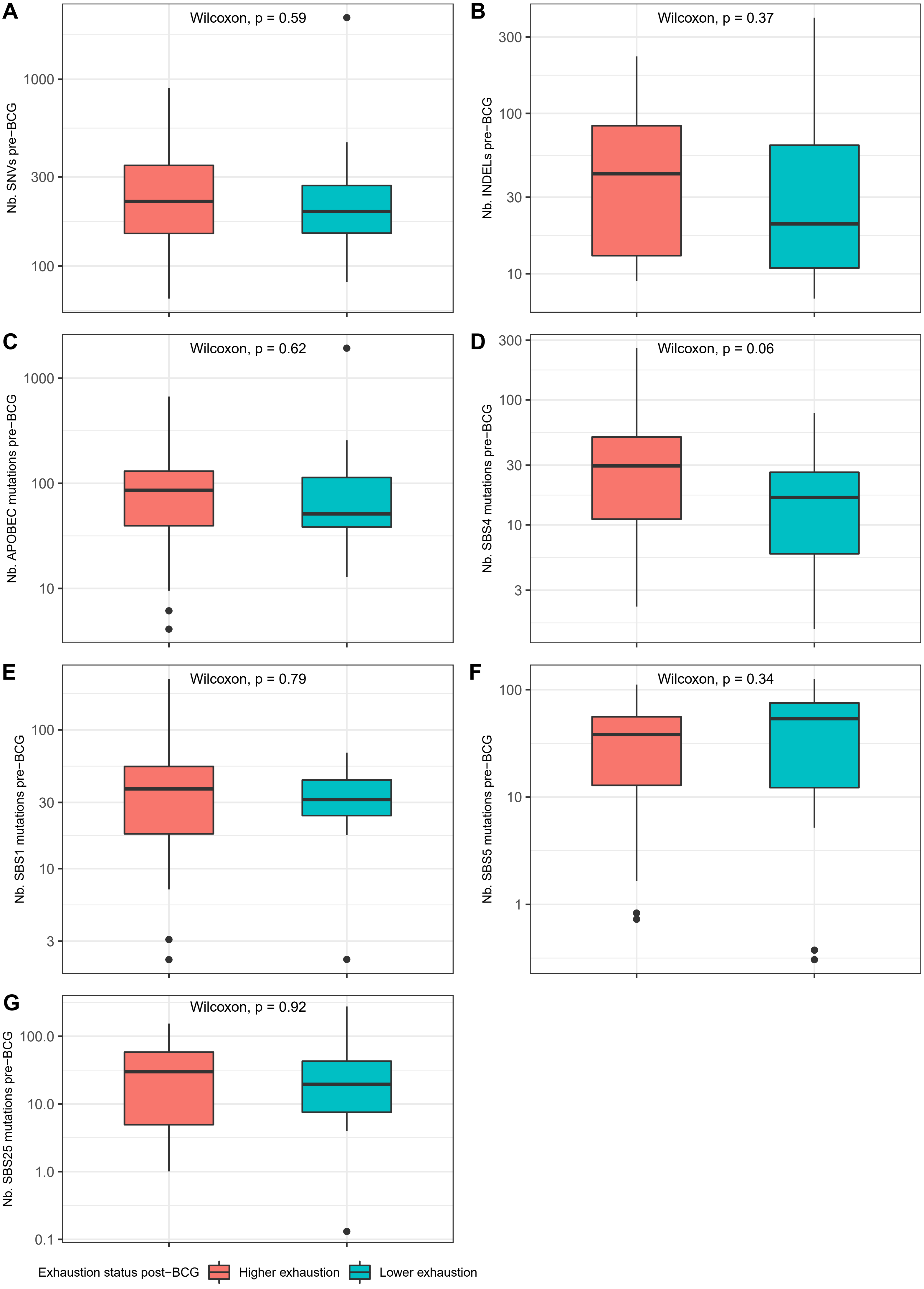

#### Supplementary Fig. S4

Correlations between post-BCG tumor exhaustion status and pre-BCG tumor WES characteristics. A) The number of single nucleotide variants (SNVs) in pre-BCG tumor samples correlated to post-BCG exhaustion status. B) The number of INDELs in pre-BCG tumor samples correlated to post-BCG exhaustion status. C) The number of APOBEC-related mutations (SBS2+SBS13) in pre-BCG tumor samples correlated to post-BCG exhaustion status. D) The number of SBS4-related mutations in pre-BCG tumor samples correlated to post-BCG exhaustion status. E) The number of SBS1-related mutations in pre-BCG tumor samples correlated to post-BCG exhaustion status. F) The number of SBS5-related mutations in pre-BCG tumor samples correlated to post-BCG exhaustion status. G) The number of SBS25-related mutations in pre-BCG tumor samples correlated to post-BCG exhaustion status. Pink = Higher post-BCG exhaustion. Turquoise = Lower post-BCG exhaustion. The Wilcoxon Rank Sum test was used for statistical comparisons.

**Supplementary Fig. S5**

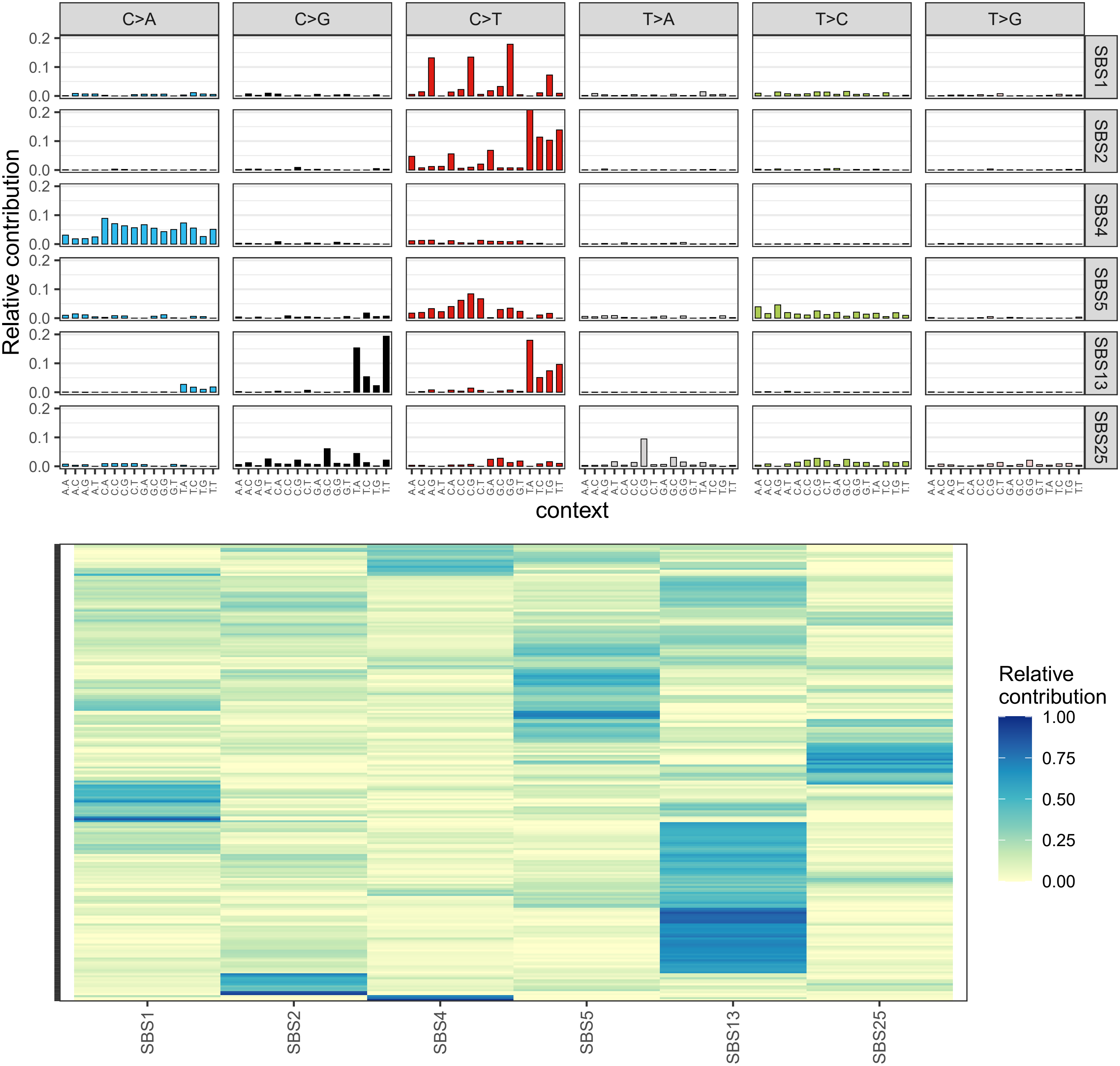

#### Supplementary Fig. S5

Overview of identified mutational signatures. A) Detailed mutational profiles for the six mutational signatures identified by de novo extraction using the R package MutationalPatterns. Extracted signatures were named according to COSMIC signatures with the highest resemblance, as estimated using cosine similarity and resulting in the following values: SBS1 = 0.91, SBS2, 0.92, SBS4 = 0.89, SBS5 = 0.75, SBS13 = 0.76, SBS25 = 0.64. B) Relative contribution of the identified mutational signatures. Sample clustering was performed based on euclidean distances.

**Supplementary Table S1:** Olink proteins and results

Overview of analyzed proteins using the Olink proteomics platform and associated biological processes. P-values and Benjamini-Hochberg adjusted P-values from paired t-tests for comparisons between pre-BCG and post-BCG samples for all patients, BCG-responsive patients, and patients with BCG-failure are shown. Green color indicates p-values < 0.05 and blue color indicates q-values <0.1.

| Olink Information | | | | All Patients | | BCG-responsive | | BCG-failure | | Test |
|---|---|---|---|---|---|---|---|---|---|---|
| *Assay* | *OlinkID* | *UniProt* | *Biological process* | *P-value* | *Adjusted p-value* | *P-value* | *Adjusted p-value* | *P-value* | *Adjusted p-value* | *Method* |
| ADA | OID00775 | P00813 | Metabolism | 2,47E-02 | 9,91E-02 | 4,12E-02 | 3,70E-01 | 1,83E-01 | 3,11E-01 | Paired t-test |
| ADGRG1 | OID00764 | Q9Y653 | Vascular and tissue remodeling | 4,50E-02 | 1,22E-01 | 8,40E-01 | 9,94E-01 | 2,38E-02 | 1,37E-01 | Paired t-test |
| ANGPT1 | OID00760 | Q15389 | Vascular and tissue remodeling | 9,39E-01 | 9,71E-01 | 6,89E-01 | 9,76E-01 | 7,24E-01 | 7,66E-01 | Paired t-test |
| ANGPT2 | OID00822 | O15123 | Vascular and tissue remodeling | 9,99E-01 | 9,99E-01 | 1,17E-01 | 5,68E-01 | 4,72E-01 | 5,37E-01 | Paired t-test |
| ARG1 | OID00815 | P05089 | Suppresses tumor immunity | 8,70E-02 | 1,96E-01 | 7,42E-01 | 9,76E-01 | 8,75E-02 | 2,24E-01 | Paired t-test |
| CAIX | OID00773 | Q16790 | Metabolism; Vascular and tissue remodeling | 2,37E-01 | 3,63E-01 | 9,32E-01 | 9,94E-01 | 1,02E-01 | 2,34E-01 | Paired t-test |
| CASP-8 | OID00827 | Q14790 | Apoptosis/cell killing | 5,17E-01 | 5,81E-01 | 7,78E-01 | 9,76E-01 | 3,83E-01 | 4,90E-01 | Paired t-test |
| CCL17 | OID00821 | Q92583 | Chemotaxis; Suppresses tumor immunity | 4,45E-01 | 5,25E-01 | 5,12E-01 | 9,01E-01 | 1,66E-01 | 3,02E-01 | Paired t-test |
| CCL19 | OID00794 | Q99731 | Chemotaxis; Suppresses tumor immunity | 6,66E-04 | 1,14E-02 | 4,27E-02 | 3,70E-01 | 7,09E-03 | 7,44E-02 | Paired t-test |
| CCL20 | OID00837 | P78556 | Chemotaxis; Suppresses tumor immunity | 3,75E-02 | 1,11E-01 | 3,40E-02 | 3,70E-01 | 1,55E-01 | 2,90E-01 | Paired t-test |
| CCL23 | OID00811 | P55773 | Chemotaxis; Vascular and tissue remodeling | 1,11E-04 | 2,55E-03 | 6,67E-03 | 1,16E-01 | 5,56E-03 | 7,31E-02 | Paired t-test |
| CCL3 | OID00813 | P10147 | Chemotaxis | 2,22E-01 | 3,53E-01 | 3,37E-01 | 7,53E-01 | 3,48E-01 | 4,78E-01 | Paired t-test |
| CCL4 | OID00796 | P13236 | Chemotaxis | 2,66E-02 | 9,91E-02 | 5,55E-02 | 4,26E-01 | 1,16E-01 | 2,38E-01 | Paired t-test |
| CD244 | OID00758 | Q9BZW8 | Promotes tumor immunity; Promotes tumor immunity | 2,84E-01 | 4,08E-01 | 8,71E-01 | 9,94E-01 | 9,24E-02 | 2,24E-01 | Paired t-test |
| CD27 | OID00800 | P26842 | Promotes tumor immunity | 2,89E-02 | 9,91E-02 | 3,12E-01 | 7,53E-01 | 5,02E-02 | 1,75E-01 | Paired t-test |
| CD28 | OID00793 | P10747 | Promotes tumor immunity | 3,30E-01 | 4,40E-01 | 7,27E-01 | 9,76E-01 | 3,41E-01 | 4,75E-01 | Paired t-test |
| CD4 | OID00776 | P01730 | Promotes tumor immunity; Suppresses tumor immunity | 6,77E-02 | 1,67E-01 | 5,19E-01 | 9,01E-01 | 5,85E-02 | 1,75E-01 | Paired t-test |
| CD40 | OID00781 | P25942 | Promotes tumor immunity | 9,46E-03 | 6,69E-02 | 2,55E-01 | 6,90E-01 | 1,64E-02 | 1,00E-01 | Paired t-test |
| CD40-L | OID00756 | P29965 | Apoptosis/cell killing; Promotes tumor immunity | 8,03E-01 | 8,49E-01 | 6,52E-01 | 9,76E-01 | 9,35E-01 | 9,45E-01 | Paired t-test |
| CD5 | OID00812 | P06127 | Promotes tumor immunity; Suppresses tumor immunity | 1,96E-03 | 1,81E-02 | 1,50E-01 | 5,87E-01 | 4,57E-03 | 7,31E-02 | Paired t-test |
| CD70 | OID00808 | P32970 | Promotes tumor immunity | 7,48E-04 | 1,14E-02 | 3,24E-01 | 7,53E-01 | 6,56E-04 | 1,51E-02 | Paired t-test |
| CD83 | OID00841 | Q01151 | Promotes tumor immunity | 2,96E-03 | 2,47E-02 | 2,13E-01 | 6,56E-01 | 5,46E-03 | 7,31E-02 | Paired t-test |
| CD8A | OID00772 | P01732 | Promotes tumor immunity | 1,60E-02 | 8,20E-02 | 1,53E-01 | 5,87E-01 | 5,55E-02 | 1,75E-01 | Paired t-test |
| CRTAM | OID00766 | O95727 | Promotes tumor immunity | 1,05E-01 | 2,25E-01 | 6,00E-01 | 9,76E-01 | 1,12E-02 | 8,33E-02 | Paired t-test |
| CSF-1 | OID00843 | P09603 | Suppresses tumor immunity | 1,66E-01 | 3,03E-01 | 8,25E-01 | 9,94E-01 | 1,06E-01 | 2,38E-01 | Paired t-test |
| CX3CL1 | OID00806 | P78423 | Chemotaxis; Promotes tumor immunity | 2,07E-02 | 9,52E-02 | 8,47E-01 | 9,94E-01 | 8,02E-03 | 7,44E-02 | Paired t-test |
| CXCL1 | OID00786 | P09341 | Chemotaxis; Suppresses tumor immunity; Vascular and tissue remodeling | 4,40E-02 | 1,22E-01 | 4,53E-01 | 8,34E-01 | 6,28E-02 | 1,75E-01 | Paired t-test |
| CXCL10 | OID00807 | P02778 | Chemotaxis; Promotes tumor immunity; Vascular and tissue remodeling | 3,12E-09 | 9,58E-08 | 6,87E-05 | 2,11E-03 | 1,23E-05 | 3,78E-04 | Paired t-test |
| CXCL11 | OID00767 | O14625 | Chemotaxis; Promotes tumor immunity; Suppresses tumor immunity; Vascular and tissue remodeling | 6,50E-11 | 3,01E-09 | 7,25E-06 | 3,33E-04 | 1,58E-06 | 1,45E-04 | Paired t-test |
| CXCL12 | OID00824 | P48061 | Chemotaxis; Suppresses tumor immunity; Vascular and tissue remodeling | 7,49E-01 | 8,01E-01 | 4,26E-01 | 8,16E-01 | 7,77E-01 | 8,12E-01 | Paired t-test |
| CXCL13 | OID00830 | O43927 | Chemotaxis; Promotes tumor immunity; Suppresses tumor immunity | 1,10E-02 | 7,24E-02 | 2,14E-01 | 6,56E-01 | 2,63E-02 | 1,42E-01 | Paired t-test |
| CXCL5 | OID00801 | P42830 | Chemotaxis; Suppresses tumor immunity; Vascular and tissue remodeling | 3,74E-01 | 4,71E-01 | 7,18E-01 | 9,76E-01 | 4,21E-01 | 5,16E-01 | Paired t-test |
| CXCL9 | OID00771 | Q07325 | Chemotaxis; Promotes tumor immunity; Vascular and tissue remodeling | 4,70E-11 | 3,01E-09 | 2,21E-06 | 2,03E-04 | 5,41E-06 | 2,49E-04 | Paired t-test |
| DCN | OID00817 | P07585 | Vascular and tissue remodeling | 1,08E-01 | 2,27E-01 | 8,83E-01 | 9,94E-01 | 5,26E-02 | 1,75E-01 | Paired t-test |
| EGF | OID00759 | P01133 | Vascular and tissue remodeling | 1,73E-01 | 3,07E-01 | 7,75E-01 | 9,76E-01 | 4,94E-02 | 1,75E-01 | Paired t-test |
| FASLG | OID00792 | P48023 | Apoptosis/cell killing | 1,26E-02 | 7,25E-02 | 1,07E-01 | 5,45E-01 | 5,29E-02 | 1,75E-01 | Paired t-test |
| FGF2 | OID00770 | P09038 | Vascular and tissue remodeling | 3,04E-02 | 9,91E-02 | 1,78E-01 | 6,18E-01 | 9,56E-02 | 2,25E-01 | Paired t-test |
| Gal-1 | OID00798 | P09382 | Suppresses tumor immunity | 6,91E-01 | 7,48E-01 | 7,27E-01 | 9,76E-01 | 4,72E-01 | 5,37E-01 | Paired t-test |
| Gal-9 | OID00779 | O00182 | Apoptosis/cell killing; Suppresses tumor immunity; Vascular and tissue remodeling | 3,81E-01 | 4,72E-01 | 8,44E-02 | 4,85E-01 | 9,01E-01 | 9,21E-01 | Paired t-test |
| GZMA | OID00804 | P12544 | Apoptosis/cell killing | 1,22E-02 | 7,25E-02 | 7,58E-03 | 1,16E-01 | 6,19E-02 | 1,75E-01 | Paired t-test |
| GZMB | OID00840 | P10144 | Apoptosis/cell killing | 2,74E-02 | 9,91E-02 | 8,41E-02 | 4,85E-01 | 1,21E-01 | 2,43E-01 | Paired t-test |
| GZMH | OID00783 | P20718 | Apoptosis/cell killing | 1,68E-01 | 3,03E-01 | 6,47E-01 | 9,76E-01 | 1,96E-01 | 3,28E-01 | Paired t-test |
| HGF | OID00803 | P14210 | Vascular and tissue remodeling | 2,99E-02 | 9,91E-02 | 3,51E-01 | 7,53E-01 | 4,35E-02 | 1,75E-01 | Paired t-test |
| HO-1 | OID00805 | P09601 | Metabolism | 9,41E-01 | 9,71E-01 | 9,46E-01 | 9,94E-01 | 9,67E-01 | 9,67E-01 | Paired t-test |
| ICOSLG | OID00828 | O75144 | Promotes tumor immunity | 3,14E-02 | 9,91E-02 | 7,51E-01 | 9,76E-01 | 1,18E-02 | 8,33E-02 | Paired t-test |
| IFN-gamma | OID05552 | P01579 | Promotes tumor immunity | 4,40E-01 | 5,25E-01 | 3,76E-01 | 7,53E-01 | 8,13E-01 | 8,40E-01 | Paired t-test |
| IL-1 alpha | OID00757 | P01583 | Promotes tumor immunity; Suppresses tumor immunity; Vascular and tissue remodeling | 2,92E-01 | 4,13E-01 | 4,08E-01 | 7,98E-01 | 4,53E-01 | 5,34E-01 | Paired t-test |
| IL10 | OID00809 | P22301 | Suppresses tumor immunity | 9,52E-01 | 9,71E-01 | 3,75E-01 | 7,53E-01 | 6,84E-01 | 7,49E-01 | Paired t-test |
| IL12 | OID00842 | P29459,P29460 | Promotes tumor immunity | 8,20E-02 | 1,93E-01 | 3,64E-01 | 7,53E-01 | 1,43E-01 | 2,79E-01 | Paired t-test |
| IL12RB1 | OID00835 | P42701 | Promotes tumor immunity | 4,48E-02 | 1,22E-01 | 2,28E-01 | 6,56E-01 | 1,13E-01 | 2,38E-01 | Paired t-test |
| IL13 | OID00836 | P35225 | Suppresses tumor immunity | 4,61E-01 | 5,36E-01 | 9,93E-01 | 9,94E-01 | 3,64E-01 | 4,78E-01 | Paired t-test |
| IL15 | OID05551 | P40933 | Chemotaxis; Promotes tumor immunity; Suppresses tumor immunity | 6,83E-01 | 7,48E-01 | 9,94E-01 | 9,94E-01 | 5,81E-01 | 6,43E-01 | Paired t-test |
| IL18 | OID00782 | Q14116 | Promotes tumor immunity; Suppresses tumor immunity | 1,56E-01 | 3,03E-01 | 2,36E-01 | 6,58E-01 | 4,06E-01 | 5,12E-01 | Paired t-test |
| IL2 | OID00778 | P60568 | Promotes tumor immunity | 3,66E-01 | 4,71E-01 | 9,78E-01 | 9,94E-01 | 2,71E-01 | 4,04E-01 | Paired t-test |
| IL33 | OID00788 | O95760 | Suppresses tumor immunity | 2,43E-01 | 3,66E-01 | 9,27E-01 | 9,94E-01 | 1,51E-01 | 2,89E-01 | Paired t-test |
| IL4 | OID00833 | P05112 | Suppresses tumor immunity | 2,75E-01 | 4,02E-01 | 6,31E-01 | 9,76E-01 | 2,99E-01 | 4,37E-01 | Paired t-test |
| IL5 | OID00802 | P05113 | Suppresses tumor immunity | 1,24E-01 | 2,47E-01 | 1,81E-01 | 6,18E-01 | 3,56E-01 | 4,78E-01 | Paired t-test |
| IL6 | OID00763 | P05231 | Promotes tumor immunity; Suppresses tumor immunity | 3,52E-01 | 4,63E-01 | 7,95E-01 | 9,76E-01 | 3,62E-01 | 4,78E-01 | Paired t-test |
| IL7 | OID00761 | P13232 | Promotes tumor immunity | 1,61E-01 | 3,03E-01 | 7,92E-01 | 9,76E-01 | 9,26E-02 | 2,24E-01 | Paired t-test |
| IL8 | OID00752 | P10145 | Chemotaxis; Suppresses tumor immunity; Vascular and tissue remodeling | 4,31E-01 | 5,22E-01 | 7,94E-01 | 9,76E-01 | 4,59E-01 | 5,35E-01 | Paired t-test |
| KIR3DL1 | OID05550 | P43629 | Suppresses tumor immunity | 3,10E-01 | 4,23E-01 | 9,78E-01 | 9,94E-01 | 1,67E-01 | 3,02E-01 | Paired t-test |
| KLRD1 | OID00839 | Q13241 | Promotes tumor immunity | 3,18E-02 | 9,91E-02 | 3,75E-01 | 7,53E-01 | 4,64E-02 | 1,75E-01 | Paired t-test |
| LAG3 | OID05553 | P18627 | Suppresses tumor immunity | 2,09E-01 | 3,44E-01 | 4,35E-01 | 8,18E-01 | 3,30E-01 | 4,68E-01 | Paired t-test |
| LAMP3 | OID00826 | Q9UQV4 | Suppresses tumor immunity | 1,37E-02 | 7,40E-02 | 1,71E-01 | 6,18E-01 | 4,19E-02 | 1,75E-01 | Paired t-test |
| LAP TGF-beta-1 | OID00785 | P01137 | Suppresses tumor immunity | 5,07E-01 | 5,81E-01 | 9,38E-01 | 9,94E-01 | 4,47E-01 | 5,34E-01 | Paired t-test |
| MCP-1 | OID00765 | P13500 | Chemotaxis; Vascular and tissue remodeling | 2,34E-01 | 3,63E-01 | 7,53E-01 | 9,76E-01 | 2,13E-01 | 3,44E-01 | Paired t-test |
| MCP-2 | OID00795 | P80075 | Chemotaxis | 1,67E-01 | 3,03E-01 | 6,66E-01 | 9,76E-01 | 1,80E-01 | 3,11E-01 | Paired t-test |
| MCP-3 | OID00755 | P80098 | Chemotaxis | 5,74E-02 | 1,47E-01 | 7,33E-02 | 4,85E-01 | 2,10E-01 | 3,44E-01 | Paired t-test |
| MCP-4 | OID00768 | Q99616 | Chemotaxis; Suppresses tumor immunity; Vascular and tissue remodeling | 3,13E-01 | 4,23E-01 | 3,52E-01 | 7,53E-01 | 5,06E-02 | 1,75E-01 | Paired t-test |
| MIC-A/B | OID00820 | Q29983,Q29980 | Suppresses tumor immunity | 5,31E-01 | 5,89E-01 | 9,46E-01 | 9,94E-01 | 4,13E-01 | 5,14E-01 | Paired t-test |
| MMP12 | OID00829 | P39900 | Suppresses tumor immunity; Vascular and tissue remodeling | 1,93E-01 | 3,26E-01 | 5,53E-01 | 9,42E-01 | 2,35E-01 | 3,73E-01 | Paired t-test |
| MMP7 | OID00814 | P09237 | Apoptosis/cell killing; Suppresses tumor immunity | 8,72E-02 | 1,96E-01 | 3,06E-01 | 7,53E-01 | 1,74E-01 | 3,08E-01 | Paired t-test |
| MUC-16 | OID05549 | Q8WXI7 | Suppresses tumor immunity | 8,67E-04 | 1,14E-02 | 3,95E-03 | 9,08E-02 | 8,07E-02 | 2,12E-01 | Paired t-test |
| NCR1 | OID00816 | O76036 | Promotes tumor immunity | 5,14E-01 | 5,81E-01 | 9,89E-01 | 9,94E-01 | 3,76E-01 | 4,87E-01 | Paired t-test |
| NOS3 | OID00777 | P29474 | Vascular and tissue remodeling | 9,61E-01 | 9,71E-01 | 1,93E-01 | 6,34E-01 | 2,51E-01 | 3,85E-01 | Paired t-test |
| PD-L1 | OID00799 | Q9NZQ7 | Suppresses tumor immunity | 9,36E-03 | 6,69E-02 | 2,25E-01 | 6,56E-01 | 1,60E-02 | 1,00E-01 | Paired t-test |
| PD-L2 | OID00831 | Q9BQ51 | Suppresses tumor immunity | 1,20E-01 | 2,45E-01 | 7,63E-01 | 9,76E-01 | 2,77E-02 | 1,42E-01 | Paired t-test |
| PDCD1 | OID00791 | Q15116 | Suppresses tumor immunity | 1,80E-03 | 1,81E-02 | 9,65E-02 | 5,22E-01 | 8,26E-03 | 7,44E-02 | Paired t-test |
| PDGF subunit B | OID00790 | P01127 | Vascular and tissue remodeling | 1,93E-01 | 3,26E-01 | 4,79E-01 | 8,64E-01 | 2,50E-01 | 3,85E-01 | Paired t-test |
| PGF | OID00762 | P49763 | Vascular and tissue remodeling | 1,95E-01 | 3,26E-01 | 9,93E-01 | 9,94E-01 | 1,10E-01 | 2,38E-01 | Paired t-test |
| PTN | OID00823 | P21246 | Vascular and tissue remodeling | 3,84E-01 | 4,72E-01 | 2,92E-01 | 7,53E-01 | 7,00E-01 | 7,49E-01 | Paired t-test |
| TIE2 | OID00754 | Q02763 | Vascular and tissue remodeling | 2,14E-01 | 3,45E-01 | 6,09E-01 | 9,76E-01 | 6,11E-02 | 1,75E-01 | Paired t-test |
| TNF | OID05554 | P01375 | Promotes tumor immunity; Suppresses tumor immunity; Vascular and tissue remodeling | 2,57E-01 | 3,81E-01 | 7,49E-01 | 9,76E-01 | 2,72E-01 | 4,04E-01 | Paired t-test |
| TNFRSF12A | OID00810 | Q9NP84 | Apoptosis/cell killing; Vascular and tissue remodeling | 1,97E-02 | 9,52E-02 | 1,45E-01 | 5,87E-01 | 7,36E-02 | 1,99E-01 | Paired t-test |
| TNFRSF21 | OID00818 | O75509 | Apoptosis/cell killing | 4,88E-02 | 1,28E-01 | 2,45E-02 | 3,22E-01 | 4,84E-01 | 5,43E-01 | Paired t-test |
| TNFRSF4 | OID00819 | P43489 | Promotes tumor immunity | 3,23E-02 | 9,91E-02 | 3,44E-01 | 7,53E-01 | 4,02E-02 | 1,75E-01 | Paired t-test |
| TNFRSF9 | OID00753 | Q07011 | Promotes tumor immunity | 2,93E-02 | 9,91E-02 | 1,44E-01 | 5,87E-01 | 1,14E-01 | 2,38E-01 | Paired t-test |
| TNFSF14 | OID00787 | O43557 | Promotes tumor immunity | 3,71E-01 | 4,71E-01 | 9,05E-01 | 9,94E-01 | 3,05E-01 | 4,38E-01 | Paired t-test |
| TRAIL | OID00769 | P50591 | Apoptosis/cell killing | 1,40E-03 | 1,62E-02 | 7,38E-02 | 4,85E-01 | 8,90E-03 | 7,44E-02 | Paired t-test |
| TWEAK | OID00789 | O43508 | Apoptosis/cell killing; Vascular and tissue remodeling | 3,10E-01 | 4,23E-01 | 1,49E-01 | 5,87E-01 | 6,95E-01 | 7,49E-01 | Paired t-test |
| VEGFA | OID00832 | P15692 | Vascular and tissue remodeling | 9,01E-02 | 1,97E-01 | 4,43E-02 | 3,70E-01 | 4,53E-01 | 5,34E-01 | Paired t-test |
| VEGFR-2 | OID00780 | P35968 | Vascular and tissue remodeling | 6,88E-02 | 1,67E-01 | 5,68E-01 | 9,50E-01 | 5,99E-02 | 1,75E-01 | Paired t-test |

**Supplementary Table S2:** Gene lists used for the RNA-based estimation of immune cell populations. Gene lists were obtained from[14], except gene markers for CD4 T-cells, which were obtained from[13].

| Immune cell population | Gene list |
|---|---|
| B cells | *MS4A1, SPIB* |
| CD4+ T cells | *AMIGO2, CD40LG, ICOS, IGFBP4, ITM2A, TRAT1* |
| CD45 | *PTPRC* |
| CD8+ T cells | *CD8A, CD8B* |
| Cytotoxic cells | *CTSW, GNLY, GZMA, GZMB, GZMH, KLRB1, KLRD1, KLRK1, NKG7, PRF1* |
| Dendritic cells | *CCL13, CD209, HSD11B1* |
| Exhausted CD8+ T cells | *CD244, EOMES, LAG3, PTGER4* |
| Macrophages | *CD163, CD68, CD84, MS4A4A* |
| Mast cells | *CPA3, HDC, MS4A2, TPSAB1, TPSB2* |
| Neutrophils | *CSF3R, FCGR3B, FPR1, SIGLEC5* |
| NK CD56- cells | *IL21R* |
| NK cells | *XCL1* |
| T cells | *CD3D, CD3E, CD3G, CD6, SH2D1A, TRAT1* |
| $T_H$1 cells | *TBX21* |
| Tregs | *FOXP3* |

### Supplementary Materials and Methods

#### Clinical samples

Tumor biopsies were either dry-frozen or fresh frozen (FF) embedded in TissueTek OCTTM Compound (Sakura Finetek), snap-frozen in liquid nitrogen and stored at -80ºC or obtained from formalin fixed paraffin embedded (FFPE) samples. Blood samples from all patients were stored in EDTA tubes at -80ºC. Urine supernatants were centrifuged and stored at -80ºC. Urine samples for protein analyses were sampled at a maximum of four months before and after BCG treatment.

#### DNA and RNA extraction

Tumor DNA from FF and dry-frozen tumors was extracted with Gentra Puregene Tissue Kit (Qiagen). DNA from FFPE samples was extracted using AllPrep DNA/RNA Kit (Qiagen). From peripheral blood, leukocyte DNA for germline (GL) reference was extracted using Qiasymphony DSP DNA midi kit (Qiagen) for all patients. Total Tumor RNA was extracted using RNeasy Mini and Micro Kits (Qiagen).

From tumors, haematoxylin and eosin (HE) stained sections of 4 µm were included to estimate carcinoma cell percentage.

DNA and RNA from tumors were extracted from serial cryosections of 20-25 µm. DNA from FFPE tumor specimens was extracted using punches taken from areas of the tumor with high estimated carcinoma cell percentage.

Urine (20–50 ml) was centrifuged at 3000G for 10 minutes, and supernatants were stored at –80C. Urine samples were thawed on ice, 10µl of urine was stored for Olink proteomics analyses (described below), PBS was added to remaining urine for a total volume of 4ml, and samples were centrifuged at 3000G for 10 minutes. Total cell-free DNA (cfDNA) was extracted from a median of 3.6 ml urine supernatants (range 0.7–3.65 ml). 10% ATL lysis buffer (median: 400µl, range: 100µl-500µl) (Qiagen, Aarhus, Denmark) was added, samples were incubated for 1.5-2h at room temperature followed by centrifugation at 1300G for 10 minutes. cfDNA extraction was performed using the QIAsymphony DSP Circulating DNA Kit (Qiagen). DNA was eluted in 60 µl ddH2O in DNA lobind tubes (Sigma-Aldrich).

#### Design of custom panel for targeted sequencing

For deep targeted sequencing of tumor specific mutations, we designed NGS panels where unique mutations (n=10-71) from 154 patients were included. Single nucleotide variants (SNVs) for the panels were selected from tumor guided WES analysis. Three panels covering mutations from 48, 54, and 52 patients, respectively, were designed.

SNVs were included based on the following criteria: 1. High or moderate impact mutations were favored. 2) Mutations with high variant allele frequencies (VAFs). 3) Mutations in known oncogenic genes from OnkoKB. 4) Bladder cancer associated genes (The Cancer Genome Atlas[1,2], MSKCC cBioPortal for Cancer Genomics[3,4], Uromol 2021[3,4], and Lamy *et al*[5]). 5) Mutations in the most exonically variable genes, reported by the Ingenuity Variant Analysis (IVA) software and/or at error-prone positions/in commonly reported erroneous contexts (C>T) were excluded, unless present in cancer driver genes or known bladder cancer genes. The three panels differed slightly in mutation selection.

#### Library preparation, deep-targeted sequencing and Mutation Calling of urinary

NGS libraries of cfDNA from urine supernatant were prepared using a modified version of the Mechanical Fragmentation Library preparation kit from Twist Bioscience in order to increase small fragment preservation and increase conversion rate. For robust error correction, 9 bp Unique Molecular Identifiers (UMI) were incorporated by replacing the twist adaptors with xGen™ UDI-UMI Adapters (Integrated DNA Technologies). The maximum cfDNA input was set to 100ng with no lower limit. Target enrichment was conducted using a modified version of the Twist Target Enrichment Protocol (Twist Bioscience) in combination with custom Twist panels (each SNV was captured by 120 bp probes on both strands). UMI consensus base calls were called using the fgbio tool package (v1.4.0). The ClipBAM (Hard-clipping) function of fgbio was enabled to avoid double count of variant calls of reads from the same template by clipping overlap between the reads. This was used for calculating UMI family size and UMI read depth. For UMI consensus collapsing, three reads were required as minimum and an editing distance of one was allowed. However, for secondary alignments, reads were not clipped. Realignment at INDEL positions were made using abra2 (v2.24). DeepSNV[6] was applied for detection and quantification of low-frequency tdDNA mutations in urine samples.

Calculations of UMI consensus read depth and UMI family sizes were based on the fgbio tool package (v1.4.0) that was run with the ClipBAM (Hard-clipping) function. Realignment at

INDEL positions are made using abra2 (v2.24). For UMI consensus collapsing, three reads were required as minimum and an editing distance of one was allowed.

Reads were mapped against the hg38 genome using bwa mem v.0.7.17. Mapped reads were grouped and consensus reads were generated from UMIs. Hereafter, read counts for the positions included in the panel were evaluated using the pileup tool bam read count. Only mutations detected in both the 5'-3' direction and the 3'-5' direction were included. Furthermore, at least three reads with different UMIs supporting a given mutation were required.

Copies per ml were calculated as follows:
GE/ml = (DNA input(ng)*VAF*(1000 pg/ng)/(3.3pg/GE))/urine (ml)
Clearance of tumor derived DNA (tdDNA) was defined as VAF ≠ 0 in pre-BCG samples and VAF=0 in post-BCG samples.

#### Neoantigen Calling

HLA types were called using POLYSOLVER[7], xHLA[8], and OptiType[9], with patient HLA type decided by consensus vote supported by at least two algorithms. If no majority could be reached, POLYSOLVER was used. All novel 9-11mer peptide fragments were generated by MuPeXI[10] and eluted ligands (EL) rank-percentage scores for all HLA alleles were predicted by NetMHCpan-4.1[11]. The rank-percentage score represents the rank of the fragments EL probability compared to a set of random natural peptides. A mutation was considered a neoantigen if at least one fragment had an EL rank percentage score<2%, for at least one HLA allele.

#### DeepSNV

The inclusion of genomic positions associated with multiple patients on every custom panel, facilitates an abundance of sequencing data for every genomic position with no mutations expected to be present. We exploited this by employing an analysis framework based on a maximum likelihood implementation of the shearwater algorithm developed by Gerstung *et al.*[6]. In brief, a background error model was built by fitting presumably non-mutated data, i.e. data from all samples not associated with a given mutation, to a binomial distribution with site-specific calculation of the dispersion for every mutation of interest. Samples with a VAF at a given position above 25% were excluded when generating the error model. Similarly, samples with a read depth below 50 at a given position were excluded when generating the error model. Furthermore, positions with an average error-rate above 10% across presumably non-mutated samples were excluded. Presumably mutated data, i.e. data from the target positions of a given sample, was assessed for a statistical significant difference compared to the background error model. Resulting *p*-values were corrected for multiple testing using the Benjamini-Hochberg procedure and adjusted *p*-values below 0.05 were considered significant.

In addition, to exploit that data were generated for all positions in a custom panel for every sample, we established an analysis to assess the combined signal from all target positions for a given sample. We used Fisher's method to calculate the sum of logs for all *p*-values obtained when analyzing the target positions. To gauge a randomly expected level of signal, we randomly sampled the same number of mutations as the target positions from non-target positions and similarly calculated the sum of logs. This was performed 10,000 times. Only samples with a sum of log scores greater than all scores from random sampling were considered ctDNA positive.

#### RNA sequencing data

Salmon[12] was used to quantify the expression of transcripts using annotation from the Gencode release 33 on genome assembly GRCh38.

Spiderplots were generated using the Radarchart function in the R package fmsb.

We estimated immune cell populations from the RNA-seq data using established gene expression signatures as in Rosenthal *et al.*[13–15]. A score for each immune cell population was calculated as the mean expression of all marker genes for the given cell type and a total immune score was defined as the mean of all immune cell population scores. Individual scores for B-cells, neutrophils and NK-cells were not evaluated as genes were missing for these cell population signatures. Gene lists are available in Supplementary Table S3.

#### WES analyses

MutationalPatterns was used for *de novo* extraction of mutational signatures[16,17]
